# Refinement of the Classification of *DDX41* Variants Through Analysis of Aggregated Clinical Datasets

**DOI:** 10.1101/2025.10.07.25335301

**Authors:** Ing Soo Tiong, Sally Hunter, Yamuna Kankanige, Nikita N. Mehta, Ryan A. Chisholm, Simon Wu, Jamilla Li, Joshua Casan, Kah Lok Chan, Lucy A. Godley, Lucy C. Fox, Piers Blombery

**Author notes:** Corresponding Author: Dr Ing Soo Tiong Department of Pathology, Peter MacCallum Cancer Centre, 350 Grattan Street, Melbourne VIC 3000, Australia E.

## Abstract

Deleterious germline *DDX41* variants are the leading cause of heritable predisposition to myelodysplastic syndrome and acute myeloid leukemia (MDS/AML). Accurate classification of pathogenicity is crucial for managing patients and their families. The absence of specific guidelines, along with late-onset disease, incomplete penetrance, and founder variants, poses challenges in clinical and laboratory practice. We aggregated a synthetic cohort (ASC) of *DDX41* germline and somatic variants from 36 studies, including 1802 cases among 53795 patients, plus an additional 832 cases from non-cohort publications. We aimed to leverage the *DDX41*-ASC to develop and refine ACMG/AMP criteria on case enrichment *(PS4*), somatic associations (*PP4*), and computational prediction (*PP3/BP4*). Analysis confirmed that deleterious germline *DDX41* variants are most common in MDS/AML. A quasi-case-control study with ancestry matching revealed overestimated odds ratios for variants in underrepresented groups. Exploiting germline–somatic associations, we developed a Bayesian multinomial model that updates the odds of pathogenicity based on the presence and number of somatic patterns. Comparison of prediction tools showed that AlphaMissense outperformed REVEL in sensitivity. These results were integrated into an online tool to facilitate the consistent application of criteria. Overall, this comprehensive analysis of *DDX41*-ASC provides an evidence framework to inform the development of *DDX41*-specific curation guidelines.

## INTRODUCTION

Deleterious germline variants in *DDX41* are the most common cause of hereditary predisposition to myelodysplastic syndromes (MDS) and acute myeloid leukemia (AML), accounting for approximately 80% of known germline predisposition cases and up to 5% of all newly diagnosed cases.^1–8^ Testing for the presence of *DDX41* variants is now standard diagnostic practice^9–11^ and included in many clinical sequencing panels used in the diagnosis of myeloid malignancies. Identification of causative germline *DDX41* variants has important clinical implications, including for diagnostic classification, disease prognosis, stem cell donor selection, prophylaxis for graft versus host disease, predictive testing of family members, and informing long-term monitoring strategies.^8–18^

The diagnosis of *DDX41*-related hematologic malignancy predisposition syndrome (MONDO 0014809) relies on accurate classification of variant pathogenicity. *DDX41* variants are typically classified into five categories: benign (B), likely benign (LB), variant of uncertain significance (VUS), likely pathogenic (LP), and pathogenic (P) according to the joint American College of Medical Genetics and Genomics and Association of Molecular Pathology (ACMG/AMP) criteria.^19^ This framework considers genomic, biological, functional, and population evidence. To further facilitate variant classification, the Clinical Genome Resource (ClinGen) provides gene-specific guidance on applying ACMG/AMP criteria. Clinical laboratories are encouraged to submit germline variant classifications to an open-source resource, ClinVar. To date, the Myeloid Malignancy Variant Expert Panel has specified curation rules for *RUNX1,*^20,21^ but no such guidelines exist yet for *DDX41*, so individual laboratories lack consensus on variant classifications.

*DDX41* presents several challenges in variant interpretation as a result of: (i) late-onset disease, (ii) incomplete penetrance, (iii) the presence of founder variants, and (iv) the absence of validated functional assays. Conversely, *DDX41* also provides opportunities for unique contributions to pathogenicity, including the specificity of somatic findings as well as a large number of published cohort studies that focus on this group of patients. Given these challenges and opportunities in variant classification, we aimed to establish an aggregated synthetic cohort of *DDX41* variants (*DDX41*-ASC) from the published literature to study refinements to variant classification. Through this *DDX41*-ASC, we aimed to examine the connection between germline *DDX41* variants and disease contexts, conduct a comprehensive quasi-case-control analysis with ancestry group matching, apply novel statistical modeling to somatic variant data, and evaluate the performance of *in silico* tools for variant effect prediction. These results will lay the foundation for developing *DDX41* curation rules that can be applied internationally to ensure consistent variant classification worldwide.

## MATERIALS/SUBJECTS AND METHODS

### Literature review

A literature review using the keyword "DDX41" on August 17, 2024, across PubMed, Medline, Web of Science, Scopus, and Embase identified 819 references, with an additional 17 through cross-referencing. Ultimately, 36 studies involving 53825 consecutive patients with hematological malignancies or cytopenias met the criteria for case-control series (**Table S1A**). Additionally, 595 cases from 55 more studies, and 250 cases from the Peter MacCallum Cancer Centre^17^ were included for data on the somatic *DDX41* variant(s) (**Table S1B**) (Supplemental Methods).

### Quasi-case-control analysis

For cases reported in the literature, the reported ethnicity was used as a proxy for genetic ancestry (**Table S1A**). Population databases (controls) used for comparison against affected individuals with germline *DDX41* included the Genome Aggregation Database (gnomAD) v4.1.0,^22^ ToMMo 54KJPN v20230626 (by the Tohoku Medical Megabank Organization),^23^ and Korean Variant Archive v2 (KOVA)^24^ (Supplemental Methods).

### Odds of pathogenicity

In principle, we could apply the method of Maierhofer et al.^3^ to infer the probability from (non-random) associations between germline and somatic variants using Bayesian reasoning. However, in practice, concern is warranted when applying this method to small samples of a specific germline variant under evaluation. Here, we developed a more stringent test for pathogenicity suitable for small sample sizes. For each germline variant, the posterior probability of observing different somatic *DDX41* variants was estimated via a multinomial distribution (Supplemental Methods). Odds of pathogenicity (OddsPath) are calculated from Tavtigian et al.’s formula,^25^ with evidence levels: very strong (≥350), strong (≥18.7), moderate (≥4.33), and supporting (≥2.08). All cases from published cohorts and our laboratory were included, regardless of diagnosis. To avoid double-counting somatic hit data from non-cohort studies, we included all cases without a somatic hit and only counted cases with unique somatic hits not reported in cohorts.

### Statistical analyses

The Fisher’s exact test was used to compare categorical variables, and the Wilcoxon or Kruskal-Wallis test was applied for numerical variables. Prevalence estimates were shown as point estimates with 95% confidence intervals (CIs). Odds ratios (OR) and 95% CIs were calculated with Haldane correction.^26^ The lower bound of the 95% CI was used to determine the strength of pathogenicity based on log_10_(2.08).^27^ Decision trees for variant classification were built using recursive partitioning (rpart package; Supplemental Methods). Receiver operating characteristic (ROC) curves were employed to compare the performance of computational predictive tools (pROC package). Lollipop plots were created using ProteinPaint^28^ with the protein domains based on Makishima et al.^2^ The analyses were performed using R version 4.4.1 (R Foundation for Statistical Computing, Vienna, Austria).

## RESULTS

### Aggregation of existing DDX41 variant literature

After excluding B and LB variants, duplicate cases, and variants with incomplete information, the existing peer-reviewed literature (see Methods) was compiled into the *DDX41*-ASC, comprising 1802 cases with *DDX41* variants from 53795 consecutive patients, along with an additional 832 cases from non-cohort studies (**Figure 1A**). A total of 451 distinct variants were identified, including 65 variants (14%) found only in non-cohort studies (**Figure 1B**). Missense variants showed the most diverse range with 261 different variants (including five with unclassified pathogenicity due to missing variant information), followed by frameshift (n=68), nonsense (n=38), and canonical splice site (n=37) variants (**Figure 1C**).

**Figure 1.**
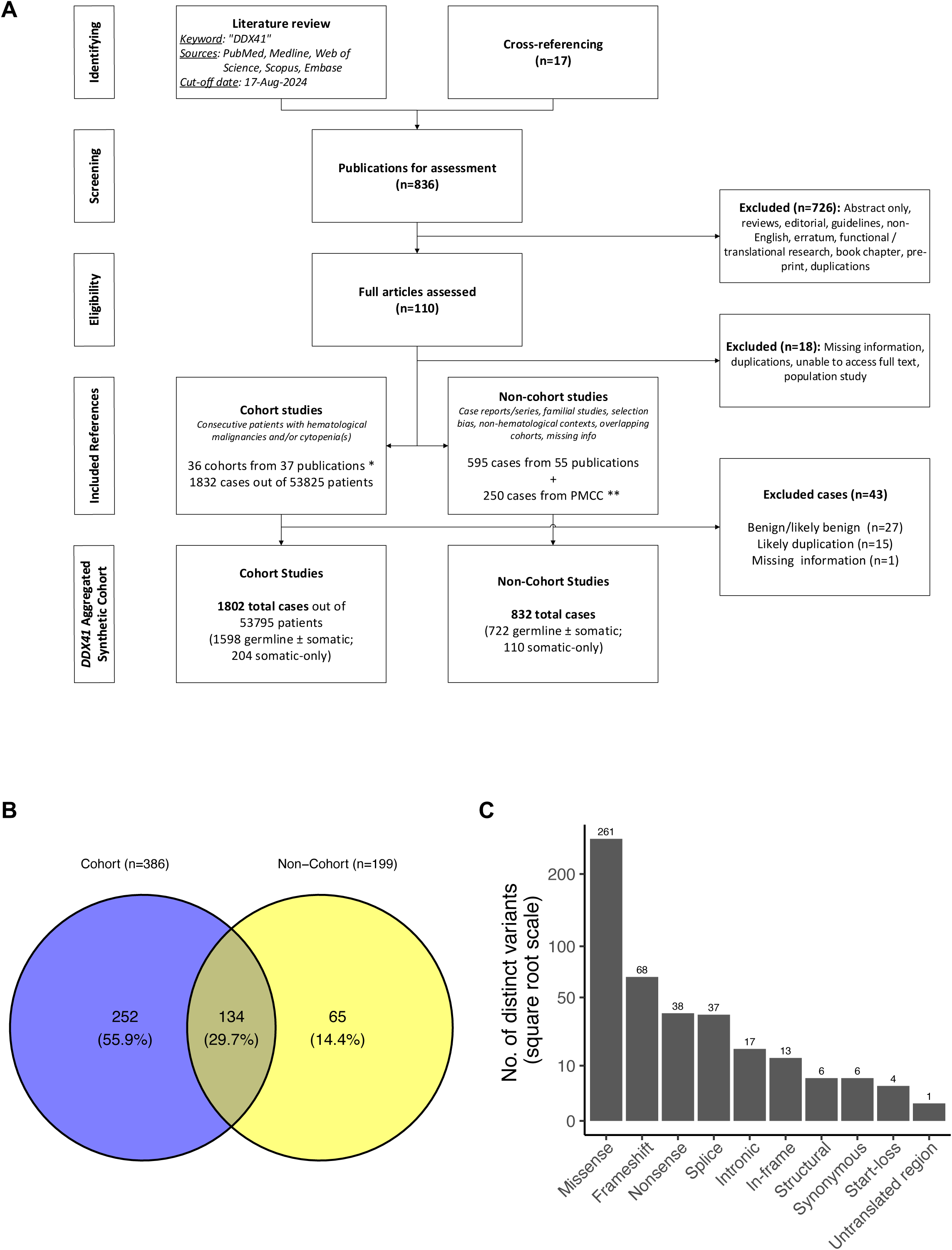
Summary of the *DDX41* aggregated synthetic cohort. **(A)** Flow diagram illustrating the literature review process for identifying published *DDX41* variants, with a cut-off date of 17-Aug-2024. *A single-center study cohort was reported in two separate publications. **Cases from the Peter MacCallum Cancer Centre (PMCC) were published in Wells et al. (2025).^17^ **(B)** Distribution of 451 distinct germline *DDX41* variants across the cohort sources. **(C)** Number of distinct *DDX41* variants by variant type.

All variants were reclassified irrespective of the classifications presented in source publications using a uniform approach (see Supplemental Methods); this is summarized and ranked using a recursive partitioning decision tree (**Figure S1**). The co-occurrence of somatic *DDX41* hotspots (evidence code *PP4*) and enrichment in MDS/AML cases (*PS4)* were key criteria contributing to classification across all variant types. Five nonsense/frameshift variants were classified as LP based on the combination of *PVS1* and absence in population controls *(PM2*_supporting).^29^ Curation of missense variants using baseline approaches resulted in most variants (n=223 [87%]) being classified as VUS.

Given the significant proportion of variants classified as VUS, we aimed to leverage the *DDX41*-ASC to develop and refine existing ACMG/AMP criteria concerning case enrichment (*PS4*), somatic associations (*PP4*), and computational prediction (*PP3/BP4*).

### Enrichment of DDX41 variants in MDS/AML (PS4)

To understand the prevalence of *DDX41* variants across different disease contexts, we included a subset of patients from the *DDX41*-ASC with a diagnosis of MDS/AML (n=34141), other myeloid neoplasms (n=8091), unexplained cytopenias (n=5156), and lymphoid neoplasms (n=1228); 5179 individuals with aplastic anemia or unspecified hematologic malignancies were excluded. Among the 1598 cases with a germline *DDX41* variant, we excluded 17 with aplastic anemia or healthy carriers, along with four cases with unclassified variants due to missing information, leaving 1577 cases included in the analysis.

Overall, a germline P/LP/VUS variant was found in 4.0% of MDS/AML cases, 2.9% of lymphoid neoplasms, 1.4% of other myeloid neoplasms, and 1.3% of cases with cytopenias (**Figure 2A**). Notably, 7 cases had a diagnosis of lymphoid neoplasm in addition to MDS/AML (n=6) or another myeloid neoplasm (n=1), and 6 MDS/AML cases had two germline variants, each being P/LP and VUS. A germline P/LP variant was found in 3.2% of MDS/AML cases (95% CI: 3.0-3.4%), which is significantly higher than in other diseases. The presence of a *DDX41* VUS was more common in lymphoid neoplasms (2%) compared to other diagnoses (0.7 to 1.0%). Among all reported germline variants, patients with MDS/AML had the highest proportion of P/LP variants (79%), followed by cases with unexplained cytopenias (45%), lymphoid neoplasms (29%), and other myeloid neoplasms (26%).

**Figure 2.**
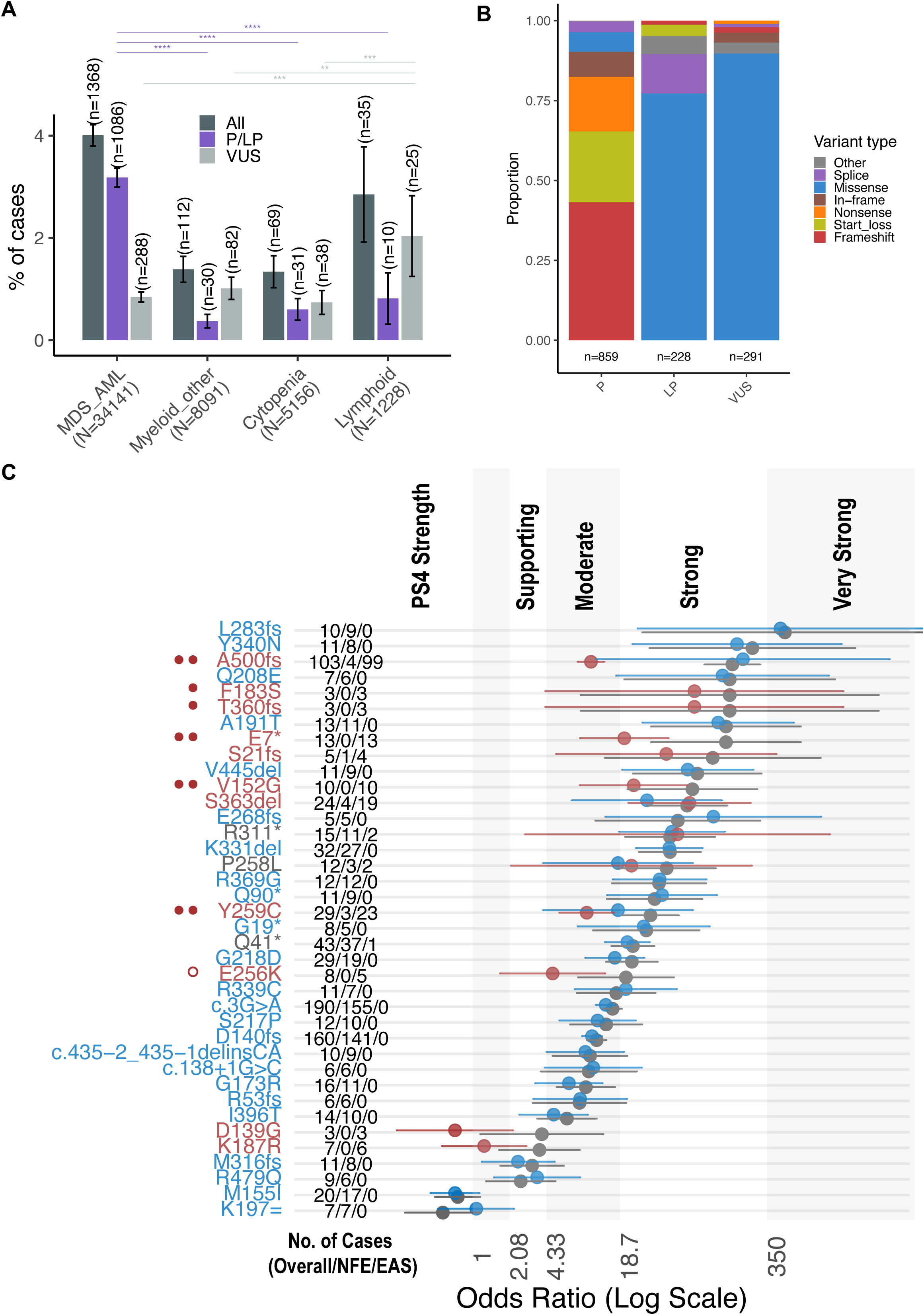
Association of germline *DDX41* variants with disease contexts and ancestral groups. **(A)** The proportion (%) of cases with pathogenic/likely pathogenic (P/LP) or uncertain (VUS) *DDX41* variants across various disease contexts in the literature cohorts. Whiskers indicate the 95% confidence interval. Twelve cases had two germline variants (6 with both VUS and P/LP were counted twice), and 7 cases had a lymphoid neoplasm in addition to either MDS/AML (n=6) or another myeloid neoplasm (n=1) (counted twice). Pairwise Fisher’s exact tests were adjusted using the Benjamini and Hochberg method. P-value annotations: <0.05 (*), <0.01 (**), <0.001 (***), <0.0001 (****). **(B)** The stacked bar chart illustrates the distribution of 1378 germline *DDX41* variants in 1368 MDS/AML cases across different classes of pathogenicity. Two cases were unclassified due to missing HGVSc information and were excluded. Ten cases harbored two germline variants: 6 were VUS + P/LP, 3 were both VUS, and 1 was both LP. Variant counts for each classification are shown below each bar. **(C)** Quasi-case-control analysis of germline *DDX41* variants in patients with MDS/AML compared to population controls, differentiated by overall and ancestral groups (non-Finnish European [NFE] versus East Asian [EAS]). Variants with at least 10 total cases, 3 occurrences within the EAS ancestry group, and 5 NFE-only instances are included. The odds ratio and its 95% confidence interval are shown. Grey, blue, and red represent overall, NFE, and EAS ancestry groups, respectively, based on the exclusivity (or lack thereof) of variants to an ancestry group and control group analyses. Red circles to the left of the variant indicate downgraded variants due to added ancestry matching: *PS4*_moderate (two solid circles), *PS4*_supporting (single solid circle), or *PS4*_notmet (open red circle).

Among 1370 MDS/AML cases, we identified 1087 P/LP, 291 VUS, and 2 unclassified (missing HGVSc) germline *DDX41* variants; 10 cases carried two germline variants (6 were VUS + P/LP, 3 were both VUS, and 1 was both LP). In total, 1378 variants corresponding to 325 distinct changes are summarized in **Figure S2**. The most common types of P variants were frameshift (43%), start-loss (22%), and nonsense (17%). Missense variants were the most prevalent variant type in LP and VUS, accounting for 77% and 90% of cases, respectively (**Figure 2B**).

Cases involving other myeloid neoplasms (myeloproliferative neoplasm [MPN; n=50], MDS/MPN [n=13], unspecified [n=48]) or unexplained cytopenias (n=69) revealed 103 distinct germline variants, with the M155I variant being the most common (8%). Notably, 70 variants occurred only once (**Figure S3A**), and 10 cases had somatic-only *DDX41* variants. There was a limited number of patients with lymphoid neoplasms across seven studies: 25 with acute lymphoblastic leukemia and 17 mature B-cell neoplasms.^1,7,28,30–33^ Of these, 35 had a germline variant, whereas seven had somatic-only *DDX41* variants (**Figure S3B**). Eight also had myeloid neoplasms: MDS/AML (n=7) and therapy-related MDS/MPN (n=1). The R164W variant, previously speculated to be associated with lymphoma,^34^ was found in three patients: lymphoplasmacytic lymphoma (LPL) with pancytopenia, gamma heavy chain disease/*MYD88*-negative LPL, and chronic lymphocytic leukemia (n=1 each).

### Ancestry group-specific variability of DDX41 variant enrichment (PS4)

After confirming the significant association between germline *DDX41* variants and MDS/AML, we conducted a quasi-case-control study of specific variants in MDS/AML cases versus population controls (see Methods). We focused on variants with at least ten cases, three occurrences in the East Asian (EAS) ancestry group, and/or five non-Finnish European (NFE)-only instances (**Figure 2C**, **Table S2**). The odds ratios for NFE-specific variants were generally consistent across overall and ancestry-specific data, except when NFE had a lower allele frequency than another group, such as for R479Q (0.03% in Admixed American versus 0.003% in NFE).

Use of gnomAD total allele counts overestimated the OR of EAS ancestry variants. As a result of added ancestry matching, the *PS4* criterion strengths were adjusted: from strong to moderate (A500fs, E7*, V152G, Y259C), moderate to supporting (F183S, T360fs), and moderate to not met (E256K) (**Figure 2C**).

Overall, eight variants showed a strong association with MDS/AML, with the lower bound of the 95% CI ≥18.7: L283fs, Y340N, Q208E, A191T, V445del, S363del, R311*, and K331del.

The two most common germline variants, c.3G>A and D140fs, were moderately enriched in MDS/AML cases (≥4.33), along with 12 other variants. Three variants met the *PS4*_supporting criterion (≥2.08).

### Characteristics of somatic DDX41 variants (PP4)

The presence of somatic *DDX41* variants is a characteristic feature in *DDX41*-related hematologic malignancy predisposition syndrome..^27^ We further characterized the observed somatic variants using the *DDX41*-ASC, which includes 34141 individuals with MDS/AML, of whom 1558 cases had a *DDX41* variant, including 830 with both germline and somatic variants, 540 with germline-only, and 188 with somatic-only *DDX41* variants.

We initially analyzed MDS/AML cases that included both germline and single somatic *DDX41* variants (n=801 cases; one excluded for missing data). As expected, R525H was the most common somatic variant (n=534; 66.7%). This was followed by six recurrent missense variants: G530D (c.1589G>A; 6.4%), P321L (c.962C>T; 4.1%), T227M (c.680C>T; 2.6%), E345D (c.1035G>C or c.1035G>T; 1.7%), G530S (c.1589G>A; 1.4%), and D344E (c.1032C>G or c.1032C>A; 1.4%) (**Figure 3A**). The remaining 15.7% of cases harbored 75 different *DDX41* variants, each with up to six instances in less than 1% of cases. Missense variants were the most common, comprising 99% of all somatic variants; six were in-frame variants, and three were truncating variants (**Figure S4A**). We observed significant differences in the median variant allele fractions (VAFs) among the somatic *DDX41* variants (**Figure S4B**): P321L (23%), T227M (13.5%), D344E (9.7%), E345D (9.5%), G530D (8.9%), R525H (7.5%), and G530S (6%).

**Figure 3.**
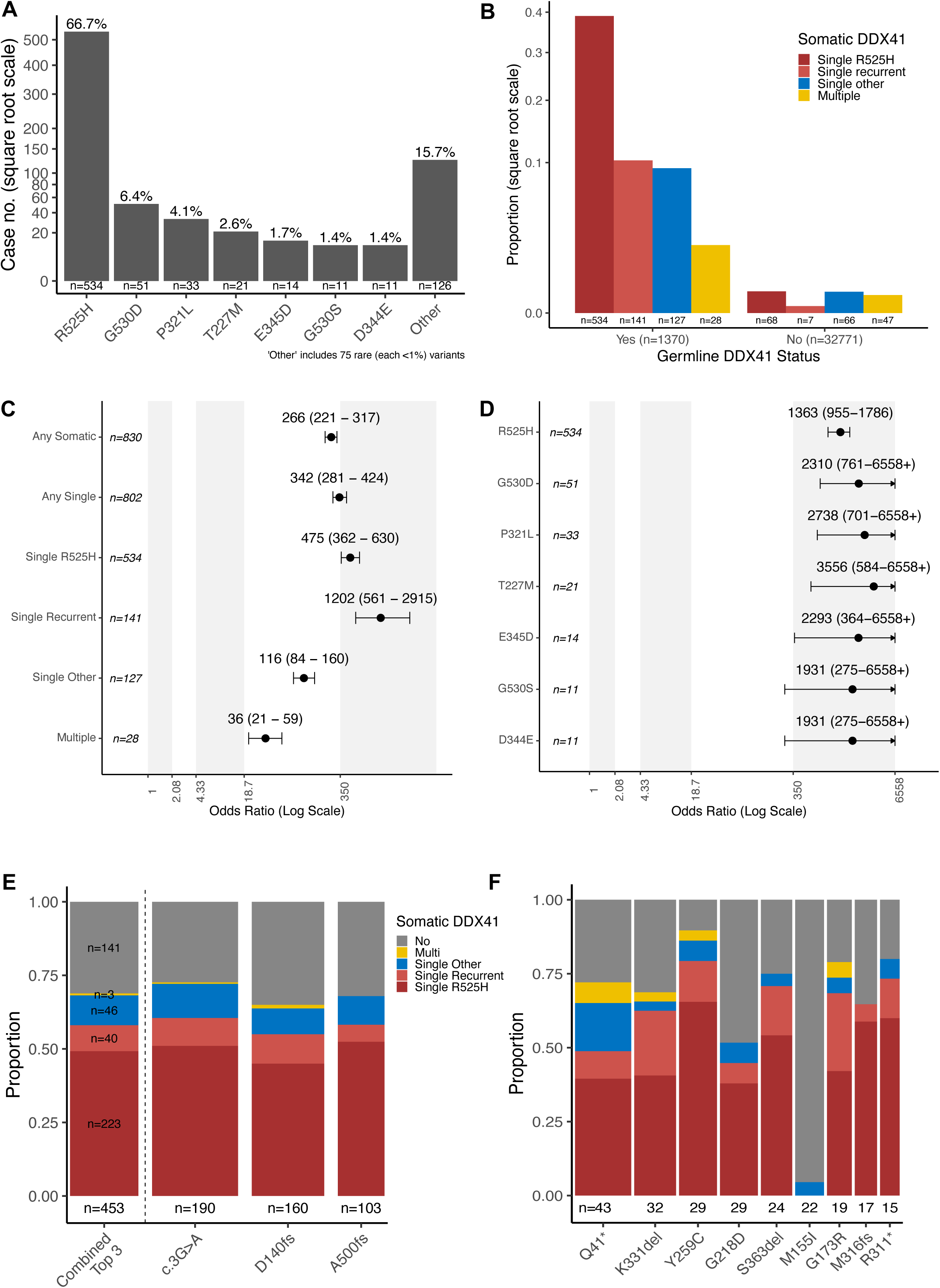
Characteristics of somatic *DDX41* variants in myelodysplastic syndrome/acute myeloid leukemia (MDS/AML). **(A)** The frequency of single *DDX41* somatic variants observed alongside a germline *DDX41* variant in 801 cases of MDS/AML. One case was excluded due to missing variant information. **(B)** The proportions of somatic *DDX41* variant types among cases with or without a germline *DDX41* variant. **(C+D)** The association between various somatic *DDX41* variants and germline *DDX41* variants in MDS/AML. Odds ratios (ORs) and 95% confidence intervals were calculated from a quasi-case-control study. Higher ORs indicate a higher prevalence of somatic variants among patients with MDS/AML who carry a germline *DDX41* variant. Case counts represent the number of cases with each somatic variant type. **(E + F)** The correlation between the most prevalent germline *DDX41* variants (occurring in at least 15 instances) and the types of somatic *DDX41* variants.

We then examined the relationship between the frequency of different types of somatic *DDX41* variants among cases with and without a germline *DDX41* variant (P, LP, and VUS). Among 1370 individuals with MDS/AML and a germline *DDX41* variant, 540 (39%) had none, 802 (58.5%) had one, 27 (2%) had two, and one had three somatic *DDX41* variants. In contrast, among the 32771 cases of MDS/AML without an identified germline *DDX41* variant, somatic *DDX41* variants were rarely found: a single variant in 141 cases (0.43%), of which 75 (0.23%) were a recurrent hotspot, and multiple variants in 47 cases (0.14%) (**Table S3**). In cases without a germline *DDX41* variant where somatic *DDX41* variants were present, they tended to be single non-recurrent or multiple variants compared to those with a germline *DDX41* variant (**Table S3**, **Figure 3B**). Indeed, a single somatic *DDX41* variant was strongly linked to a germline P/LP/VUS *DDX41* variant (OR = 342, 95% CI: 281–424) **(Figure 3C)**. This association was even stronger when R525H was considered alone (OR = 475, 95% CI: 362–630) or only recurrent non-R525H (OR = 1202, 95% CI: 561–2915) somatic variants **(Figure 3D)**. Other single non-recurrent or multiple somatic variants remained significantly associated with a germline variant, though to a lesser degree (**Figure 3C**).

The association between recurrent somatic *DDX41* variants was consistent across the well-established P/LP germline DDX41 variants (**Figure 3E, 3F**). Among the three most frequent germline *DDX41* variants—c.3G>A, D140fs, and A500fs (combined n=453)—a single somatic R525H variant was found in 45–52% of cases, single recurrent non-R525H missense variant in 6–10%, single non-recurrent variant in 9–12%, and multiple somatic variants in 0–1% (**Figure 3E**). This pattern is similarly observed for other less common but recurring germline P/LP *DDX41* variants (**Figure 3F**). In contrast, the M155I variant (classified as a VUS; gnomAD frequency 0.04%) was observed only once with a single non-recurrent somatic *DDX41* variant. The association with other less common germline variants is summarized in **Figure S5**.

The variant details of 188 cases of MDS/AML with somatic-only *DDX41* variants are summarized in **Figure S6**. Among 141 cases with a somatic-only DDX41 variant, 91% were missense, but recurrent non-R525H missense variants were rare. One case had three somatic-only *DDX41* variants: L87V, Y451C, and G586R. The remaining 46 cases had two somatic-only variants: one missense (R525H in 67%) or in-frame, combined with either another missense/in-frame or truncating variant, with both combinations occurring equally (**Figure S6**). When seen as single or double variants, the VAFs of (assumed) somatic-only *DDX41* variants were similar to those with a germline variant (**Figure S7**).

Overall, these findings support a specific association between deleterious *DDX41* variants and the pattern of somatic second hits. In contrast, although somatic-only *DDX41* variants can occur, they are much rarer and have different variant profiles.

### Odds of pathogenicity from somatic DDX41 variants (PP4)

After observing a strong non-random association between germline and somatic *DDX41* variants and building on previous work,^3^ we sought to evaluate the pathogenicity of germline *DDX41* variants informed by the presence, number, and pattern of somatic variants. In cases of MDS/AML, the three most common pathogenic variants (c.3G>A, D140fs, and A500fs) were observed to have single recurrent missense (both R525H and non-R525H), single non-recurrent, and multiple somatic variants in 58%, 10%, and 0.7% of cases, respectively (**Figure 3E**). In contrast, these somatic patterns were observed in 0.23%, 0.20%, and 0.14% of cases without a germline *DDX41* variant (**Figure 3B**). We calculated the posterior probability of pathogenicity and OddsPath^35^ using a multinomial probability mass function, noting that an observation of an isolated case with a recurrent somatic hit has a posterior probability of 97% and an OddsPath of 252 (**Figure 4A**), equivalent to a “strong” level of evidence for pathogenicity in the modifications suggested to the ACMG/AMP evidence framework.^35^

**Figure 4.**
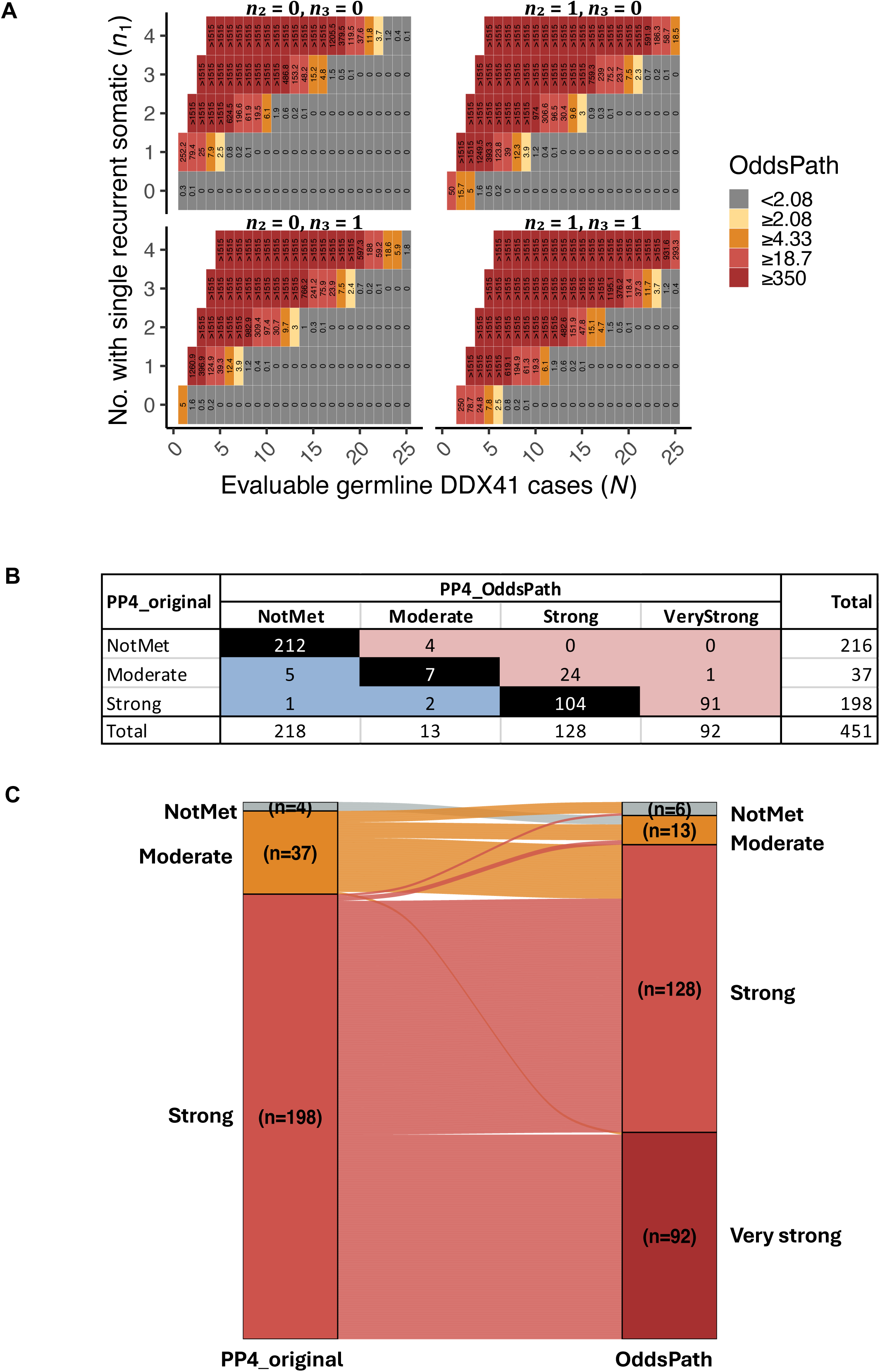
Odds of pathogenicity (OddsPath) based on the multinomial probability distribution of somatic *DDX41* variants. **(A)** Simulation of OddsPath based on the observed counts of single recurrent (R525H and non-R525H) somatic missense variants (*n*_1_), single non-recurrent somatic variants (*n*_2_), and multiple somatic hits (*n*_3_) among up to 25 evaluable cases (*N*) of germline *DDX41* variants. **(B + C)** Contingency table and Sankey diagram comparing evidence strength levels between the original approach (modified from Maierhofer et al. 2023)^41^ and OddsPath. In the Sankey diagram, 212 cases that do not meet both criteria are excluded.

Details of somatic occurrences of all 451 distinct germline *DDX41* variants are provided in **Table S4**. Overall, 92 variants had an OddsPath ≥350, consistent with a “very strong” level of evidence (**Figure 4B, 4C**). Twenty-five variants were upgraded from *PP4*_moderate to *PP4*_strong (n=24) or very strong (n=1): five from recognizing additional somatic hotspots, and the remaining from non-recurrent single or multiple somatic hits.

In contrast, eight variants had evidence downgraded based on OddsPath (**Table S4**). These included six with OddsPath <2.08: M155I (n=39), K187R (n=19), R219H (n=12), R339L (n=6), R525H (n=5), and P321L (n=5). For R525H or P321L, only two of five cases each had confirmed germline origin. Two variants (I207T and c.138+5G>T) were downgraded from *PP4*_strong to *PP4*_moderate because only one somatic hotspot was observed out of four.

When calculating the OddsPath, it is essential to consider all evidence of somatic occurrences. Incorporating non-cohort cases resulted in a total of 55 upgrades, including 40 variants from an OddsPath of <2.08, to *PP4*_moderate (n=1), strong (n=33), and very strong (n=6).

### In silico tool comparison for missense variants (PP3/BP4)

Given the numerous missense *DDX41* variants classified as VUS, we assessed the REVEL score^35^ for missense variants and compared it with AlphaMissense.^35^ After removing the *PP3* criterion, only 21 missense variants remained as P/LP. Therefore, we used all germline missense variants co-occurring with a single recurrent somatic hit to create a pathogenic truth set (n=61), excluding those within the splice junctions. We retrieved 678 missense variants from gnomAD v4.1.0. After curation, only four were LB, so we included 503 in the benign truth set, excluding 171 found in the *DDX41*-ASC.

Using the REVEL score, the receiver operating characteristic (ROC) curve demonstrated an area under the curve (AUC) of 0.79 (95% CI: 0.74–0.84), and the optimal REVEL score threshold (Youden’s index) was 0.33 (**Figure 5A**). The performance of three REVEL thresholds (≥0.33, 0.64^36^ and 0.70^3^) was compared in **Table S5**.

**Figure 5.**
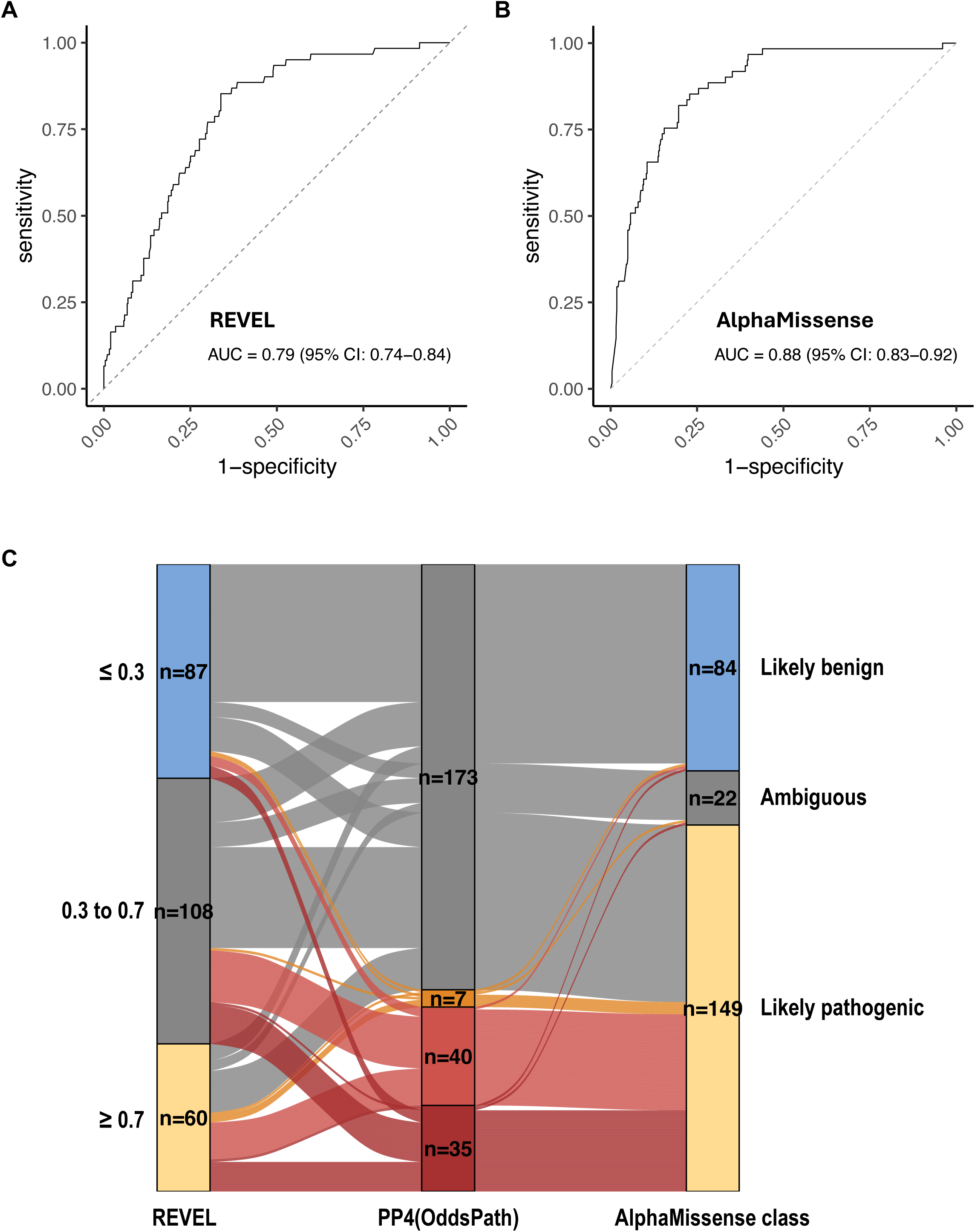
Comparison of REVEL and AlphaMissense *in silico* tools. The ability to classify putative pathogenic (n=61) and non-pathogenic (n=503) variants, based on the presence of any concurrent single recurrent somatic variant, was evaluated and compared. **(A)** Receiver Operating Characteristic (ROC) curve based on REVEL scores. **(B)** ROC curve based on AlphaMissense scores. **(C)** Sankey diagram illustrating the classification of variants as pathogenic supporting, benign supporting, or not met, based on REVEL scores (≥0.7 and ≤0.3) and AlphaMissense class, across 255 evaluable missense variants with varying levels of odds of pathogenicity (OddsPath, based on the presence of somatic *DDX41* variant). Six missense variants are excluded due to missing HGVSc information (n=5) or delins variant type (n=1). Note that this is not the final *PP3* or *BP4* classification, which also considers the potential splicing impact (e.g., by SpliceAI score).

We then evaluated AlphaMissense’s ability to identify putative pathogenic variants. Alpha scores outperformed REVEL with an AUC of 0.88 (95% CI: 0.83–0.92; p<0.001 by Delong test) (**Figure 5B**). However, we chose the pre-calculated alpha class for further analysis for its higher sensitivity (**Table S5**). REVEL was better at identifying non-pathogenic variants, though there was significant overlap. Conversely, putative pathogenic variants clustered around high alpha scores (**Figure S8**). Applied to *DDX41*-ASC (255 evaluable variants), AlphaMissense better identified variants with higher OddsPath (based on multinomial somatic hits), but had more false positives (**Figure 5C**). Of 82 variants with OddsPath ≥4.33, 77 (94%) and 32 (39%) met *PP3* by AlphaMissense and REVEL. Of 173 variants with OddsPath <2.08, 81 (47%) and 76 (44%) met *BP4* by AlphaMissense and REVEL, respectively, while 72 (42%) and 28 (16%) met *PP3*.

### Updated DDX41 variant classification

Finally, we integrated the above analysis to classify 439 evaluable germline *DDX41* variants (**Figure 6**). A total of 65 variants were upgraded: 35 from VUS (26 to LP and 9 to P), and 30 from LP to P. Remarkably, these included 33 missense variants initially classified as VUS, based on a combination of high OddsPath from observed somatic hits (*PP4*), supporting-to-moderate level of case enrichment (*PS4*) in MDS/AML, and predicted deleterious effects by AlphaMissense (*PP3*). Thirty-one variants with high *PP4*_OddsPath (8 moderate, 21 strong, and 2 very strong) remained classified as VUS, particularly affecting missense (n=20) and in-frame (n=5) variants, due to the lack of other applicable criteria.

**Figure 6.**
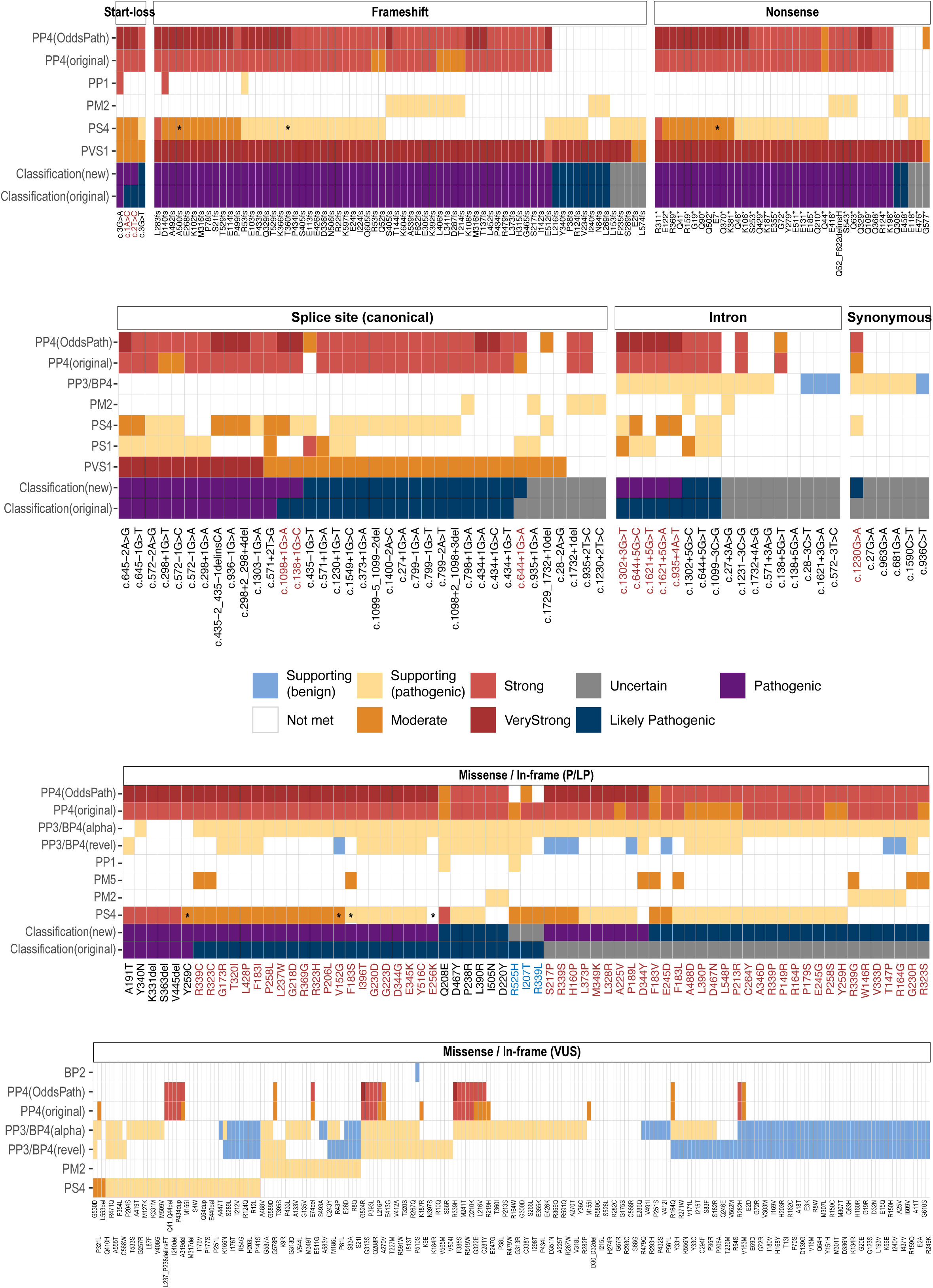
Summary of *DDX41* variant curation based on modified ACMG/AMP criteria. Each of the 439 variants is shown on the x-axis (using abbreviated nomenclature), including start-loss (n=4), frameshift (n=68), nonsense (n=38), canonical splice site (n=37), intronic (n=17), synonymous (n=6), missense (n=66) or in-frame (n=3) pathogenic / likely pathogenic (P/LP), and missense (n=190) or in-frame (n=10) variants of uncertain significance (VUS). Twelve variants were excluded: structural (n=6), untranslated region (n=1), and missing variant information (n=5 missense variants). The applicable ACMG/AMP criteria are shown in each row, including the comparison between two *PP4* approaches (modified from Maierhofer et al.^3^ [PP4(original)] versus odds of pathogenicity from multinomial probability [PP4(OddsPath)]) and *PP3/BP4* approaches (REVEL [PP3/BP4(revel)] versus AlphaMissense [PP3/BP4(alpha)]). The asterisks (*) on *PS4* indicate the revised strength of evidence based on matching for East Asian genetic ancestry. Comparisons are made between the original and updated pathogenicity classifications, with red and blue text indicating upgraded and downgraded variants, respectively. Note that five variants have two different HGVSc descriptions (listed from left to right in the order of appearance) and are shown twice: R53fs (c.155dup, c.156_157insA); M316fs (c.947_948del, c.946_947del); T529fs (c.1585dup, c.1586_1587del); G72R (c.214G>A, c.214G>C); and M155I (c.465G>C, c.465G>A).

We created an automated application that interfaces with the *DDX41*-ASC to support classification of pathogenicity according to ACMG/AMP criteria (https://blombery-lab.shinyapps.io/ddx41). In addition to calculating the odds ratio for case enrichment and OddsPath based on observed somatic hits, the application also features a customizable user interface, allowing users to specify disease contexts, second somatic hotspots, *in silico* tools for *PP3/BP4* (REVEL or AlphaMissense), and thresholds for various curation criteria such as *PS4*, REVEL,^37^ SpliceAI,^38^ and population allele frequency. Each queried variant provides detailed information, including relevant literature and related somatic hits. Users can also manually override the criteria for pathogenicity classification. The application supports bulk curation of variants, enabling multiple variants to be curated simultaneously with the standardized application of pre-specified rules.

## DISCUSSION

Evidence-based and reproducible classification of *DDX41* variants by clinical molecular pathology laboratories/services and researchers is critical for optimal patient management. To this end, we have comprehensively aggregated existing published data into a large synthetic cohort comprising 54627 total patients, 2634 total germline and somatic *DDX41* cases, and 451 unique germline *DDX41* variants. The *DDX41*-ASC has enabled analyses that have provided insights informing the evidence of pathogenicity as per existing variant curation frameworks (ACMG/AMP).

A hallmark of cancer predisposition genes is the significant enrichment in patients with a given phenotype compared to matched controls. Given the relatively high frequency of *DDX41* variants in the population due to their minimal impact on reproductive fitness, very large case-control studies are required to study disease associations effectively. Although several large MDS/AML cohorts have been documented in the literature, gathering sufficient evidence to meet this criterion (*PS4*) requires labor-intensive manual review of publications. Furthermore, because founder variants are more common in certain ancestry groups, unadjusted case-control comparisons may overestimate enrichment. The creation of a *DDX41*-ASC enables the reproducible assessment of *DDX41* variants with improved statistical accuracy.

One key characteristic of myeloid malignancy in the context of *DDX41*-related hematologic malignancy predisposition syndrome is the presence of a second somatic variant in *DDX41*. Several groups have relied on the presence of either recurrent (variously defined) or any somatic hit to inform variant pathogenicity.^1,8,39,40^ Our previous work demonstrated a highly non-random co-occurrence between deleterious germline and somatic *DDX41* variants (posterior probability 99.8%), suggesting that such findings could help strengthen the *PP4* criterion to a very strong level.^3^ In this current work, we refined the approach by incorporating the frequency with which a germline variant is observed alongside different (multinomial) somatic patterns. This method addresses the inevitable issue of somatic *DDX41* variants being coincidentally observed with a given germline *DDX41* variant, thereby preventing a false attribution of pathogenicity to the germline variant. Our proposed multinomial statistical model enables dynamic updating of the posterior probability and OddsPath, while reducing the chance of random co-occurrence.

The use of computational (*in silico*) prediction tools, although mainly providing minor evidence of pathogenicity, can still significantly impact the final variant classification. Our analysis revealed that the commonly used REVEL score thresholds lacked sufficient sensitivity for identifying putative pathogenic missense variants in *DDX41*, resulting in under-classification in many instances. In contrast, AlphaMissense—a newer deep learning-based model—demonstrated superior diagnostic performance. Implementing AlphaMissense caused a significant shift in both *PP3* and *BP4* calls, with more variants reaching the threshold for *PP3*. This increased sensitivity did not compromise specificity when combined with other criteria, as no putative benign variants (lacking recurrent somatic hits) were incorrectly classified as P/LP.

Our work has several important limitations to acknowledge. The absence of detailed ancestry information in many source publications hampers more precise quasi-case-control analysis. Most available data come from individuals of NFE and EAS ancestries, which limits the generalizability of our findings to other populations and highlights the need for more data from diverse ancestry groups. The variant and clinical data were manually extracted from various publications that vary in nomenclature, sequencing platforms, bioinformatic pipelines, and reporting standards. Therefore, intronic and structural variant types are likely underrepresented. Finally, most publications assumed the germline versus somatic origin of the *DDX41* variants rather than basing this assumption on direct evidence from paired testing with non-hematological tissue.

In summary, by creating a *DDX41*-ASC and related analyses, we have made multiple refinements in variant classification. These include an ancestry-matched quasi-case-control study for a more precise assessment of case enrichment, enhanced sophistication in incorporating *DDX41* somatic variants into classification, and the identification of AlphaMissense as a potentially preferred computational tool for pathogenicity assessment over REVEL. Ongoing collaboration and data sharing will be essential to refine these recommendations further and help incorporate them into standard diagnostic practice, facilitated by a publicly available online tool. We look forward to the incorporation of our work into ClinGen-approved *DDX41* variant curation rules, which can be implemented broadly in clinical laboratories worldwide.

## Data Availability

All data produced in the present study are available upon reasonable request to the authors

https://github.com/blomberylab/ddx41

## Acknowledgements

The authors gratefully acknowledge funding sources including the Wilson Centre for Blood Cancer Genomics and the Snowdome Foundation.

## Author Contributions

IST and PB designed the study. IST, SH, SW collected data. YK developed the online curation application. IST, SH, YK, NNM, RAC, JL, JC, KLC, LAG, LCF, PB analyzed and interpreted data. IST wrote the first draft of the manuscript. All authors reviewed and approved the final version of the manuscript.

## Data Availability Statement

Code available at https://github.com/blomberylab/ddx41

