## Supplemental Methods for "Refinement of the Classification of *DDX41* Variants Through Analysis of Aggregated Clinical Datasets"

**Additional supplemental files:**

- Supplemental Tables
- Supplemental Figures

**SupplementaL methods**

***Data extraction from the literature review***

*DDX41* variants were consistently reported using NM_016222.4. Each variant was extracted using HGVSc nomenclature whenever possible; if unavailable, back translation from HGVSp to a unique single-nucleotide variant was attempted using Mutalyzer (v3.1.1).^1^ Cases lacking HGVSc are assumed to share the same HGVSc if only one is available for a given HGVSp. For start-lost variants (p.M1?), it is assumed to be c.3G>A when no HGVSc is provided. If a germline variant was observed only twice and associated with the same non-R525H somatic variant, the second was excluded to prevent potential duplication of cases published from the same site. Variants were grouped based on their reported (confirmed or assumed) germline and somatic status; if not specified, those with a variant allele frequency (VAF) >35% for insertions/deletions and >40% for single-nucleotide substitutions were assumed to be of germline origin. Variants are categorized as pathogenic (P), likely pathogenic (LP), and variant of uncertain significance (VUS) according to modified ACMG/AMP criteria. Additional variables extracted from the publications, where available, include ethnicity/country of origin, diagnosis (myelodysplastic syndrome or acute myeloid leukemia [MDS/AML], other myeloid neoplasms, cytopenia, lymphoid neoplasms, and other), and variant allele fraction (VAF, %).

### ***Recursive partitioning***

A recursive partitioning decision tree was generated using the rpart (version 4.1.23) and rpart.plot (version 3.1.2) packages, with modified ACMG/AMP criteria (see below) as the explanatory variables and the final variant classifications as the outcome variable. The following settings were used: a complexity parameter of 0.01, a minimum split of 5, and a maximum depth of 4.

### ***Quasi-case-control analysis***

For cases reported in the literature, the reported ethnicity was used as a proxy for genetic ancestry. When ethnicity was not specified, it was inferred based on the country where the study centers were located. In this analysis, only non-Finnish European (NFE) and East Asian (EAS) ancestry groups and MDS/AML diagnoses were included. Thai was grouped with Chinese, Japanese, and Korean as part of the EAS ancestry group. Nine references were excluded for the following reasons: six did not provide specific information on the number of cases diagnosed with MDS/AML among other reported diagnoses; two did not report the ethnicity of cases with *DDX41* variants; and one consisted of individuals of Finnish origin. We classified 17 publications as NFE ancestry: eight from Europe, seven from the USA, one from Australia, and one from the Center for International Blood and Marrow Transplant Research (CIBMTR). Of the remaining publications, eight were classified as EAS ancestry (three from China, three from Korea, one from Japan, and one from Thailand), and two were mixed (the denominator is weighted according to the information provided in the manuscripts) (Table S1A).

Population databases (controls) used for comparison against affected individuals with *DDX41* included the Genome Aggregation Database (gnomAD) v4.1.0,^2^ ToMMo 54KJPN v20230626 (by the Tohoku Medical Megabank Organization),^3^ and Korean Variant Archive v2 (KOVA).^4^ Genetic ancestry groups were as reported in gnomAD, and were assumed to be EAS in ToMMo and KOVA databases. Total and NFE-specific allele numbers and counts were obtained from gnomAD v4.1.0. For the EAS ancestry group, allele numbers and counts were combined from all three databases.

### ***DDX41 curation using modified ACMG/AMP criteria***

**PVS1_variable:** Applied as per Abou Tayoun et al. (2018).^5^

**PS1_variable:** Applied for missense variants per the original ACMG/AMP guidelines,^6^ and for splice site variants per Walker et al. (2023).^7^

**PS4_variable:** The odds ratio was calculated by comparing the frequency of all patients with the variant and MDS/AML reported in case-control series in the literature to all individuals with the variant in gnomAD v4.1.0 as the control group. Various strength levels were applied based on the lower bound of the 95% confidence interval being ≥350 (2.08^8; very strong), 18.7 (2.08^4; strong), 4.33 (2.08^2; moderate), and 2.08 (supporting), respectively.^8-10^

Additionally, in this study, the above was calculated using reported or assumed ethnicity as a proxy for genetic ancestry groups. The revised PS4 evidence strengths were then used for updating the variant classification.

**PM2_supporting:** Applied if the variant is absent from gnomAD v4.1.0 and does not meet the PS4 criterion.

**PM5:** Per the original ACMG/AMP guidelines.^6^ No missense variants met this criterion at baseline curation.

**PP1_variable:** Application of PP1 at different strength levels was based on the number of segregations and number of families with the variant and MDS/AML, as directed per Jarvik and Browning (2016).^11^

**PP3/BP4:** REVEL scores of ≥0.7 and ≤0.3 for a missense variant resulted in applying PP3 and BP4, respectively. SpliceAI delta scores of ≥0.2 and ≤0.1, based on hg38 and 10,000 flanking nucleotides, result in the application of PP3 and BP4 for intronic (outside +/- 1 and +/- 2), synonymous, and/or missense variants.

Additionally, in this study, the updated PP3 and BP4, based on the AlphaMissense class (likely pathogenic and likely benign), were used to inform the final variant classification.

**PP4_variable:** According to Maierhofer et al. (2023),^12^ with some modifications. PP4_strong was applied if the germline *DDX41* variant was observed with any single (assumed) somatic *DDX41* variants in R525H, G530D, G530S, P321L, or T227M, and PP4_moderate for any other single (assumed) somatic *DDX41* variants, considering all disease contexts. This was not applicable when multiple (assumed) somatic *DDX41* variants were observed.

In this study, new PP4 strengths based on multinomial probability and odds of pathogenicity (OddsPath) were used to update the variant classification.

**BA1/BS1:** We used the Whiffin/Ware calculator (http:cardiodb.org/allelefrequencyapp/) with the following estimates: prevalence of AML of 1 in 3650 (from SEER database 2023), penetrance 50% (from Makishima et al., 2023),^13^ an allelic heterogeneity of 100%, and a maximum genetic heterogeneity of 100%, based on a 95% confidence interval. The calculated maximum credible population allele frequency, 0.000274. For BS1 and BA1, the value was increased by 10- and 100-fold, to provide the threshold of ≥0.00274 (0.274%) and ≥0.0274 (2.74%), respectively.

**BP2:** Applied when a variant was found in homozygosity in gnomAD v4.1.0. Not applied if observed with a somatic hotspot variant.

**BP7:** Applied for synonymous and intronic variants outside donor and acceptor splice regions when SpliceAI delta scores ≤0.1, without considering the evolutionary conservation score, per Walker et al. (2023).^7^

Other criteria were not applicable either in the context of the gene or for the specific variants. When the evidence for pathogenic and benign is conflicting, the Bayesian point system is used.^8,9^

### ***Bayesian posterior using multinomial distribution***

Our goal is to infer the probability that a germline *DDX41* variant is pathogenic, based on a relatively small group of patients with the variant and data on any somatic variants they may carry. In principle, we could apply the method of Maierhofer et al. (2023), which infers such a probability from (non-random) associations between germline and somatic variants using Bayesian reasoning. However, in practice, concern is warranted when applying this method to small samples of a specific germline variant under evaluation. Moreover, updating the posterior probability is impractical because the prior data (Table S3) is much larger than the new variant under study (often fewer than 10).

Here, we apply a test for pathogenicity that is more stringent and therefore may be more applicable to small sample sizes. We test whether frequencies of somatic variants associated with germline variants are not only non-random but also consistent with the frequencies observed in known pathogenic germline variants. Our method here then involves a standard application of Bayes’ rule to generate posterior probabilities of the germline variant being pathogenic.

Given a sample of patients with a germline variant and an observed frequency distribution of somatic variants among these patients, we would like to quantify the probability that the germline variant is pathogenic by comparing this frequency distribution to those from patients with known pathogenic germline variants and those without. Starting from Bayes’ rule, we have

$$\begin{aligned} \Pr\left( A | B \right)= \frac{\Pr\left( B | A \right)\Pr\left( A \right)}{\Pr\left( B \right)}\#\left( 1 \right) \end{aligned}$$

where in our problem the event $A$ is the event of a germline variant being pathogenic, and event $B$ is the observed somatic variant frequency distribution for patients with this germline variant. We can expand the denominator in Bayes’ rule as follows using the law of total probability:

$$\begin{aligned} \Pr\left( A | B \right)= \frac{\Pr\left( B | A \right)\Pr\left( A \right)}{\Pr\left( B | A \right)\Pr\left( A \right)+\Pr\left( B | A^{'} \right)\Pr\left( A^{'} \right)}\#\left( 2 \right) \end{aligned}$$

Let the frequency of somatic variants in the presence of a pathogenic germline variant be $p$ and in the absence of one be $q$, and let the sample size for our novel germline variant be $N$ and the observed number of somatic variants in our sample be $n$. Then under the binomial distribution we have

$$\begin{aligned} \Pr\left( B | A \right)=\left( \begin{matrix} N \\ n \end{matrix} \right)p^{n}\left( 1-p \right)^{N-n}\#\left( 3 \right) \end{aligned}$$

and

$$\begin{aligned} \Pr\left( B | A^{'} \right)=\left( \begin{matrix} N \\ n \end{matrix} \right)q^{n}\left( 1-q \right)^{N-n}\#\left( 4 \right) \end{aligned}$$

This could be further extended to the four types of mutually exclusive outcomes: single recurrent somatic ($n_{1}$), single other somatic ($n_{2}$), multiple somatic hits ($n_{3}$), and no somatic hits ($N-n_{1}-n_{2}-n_{3}$).

$$\begin{aligned} \Pr\left( B | A \right)=\frac{N!}{n_{1}!n_{2}!n_{3}!(N-n_{1}-n_{2}-n_{3})!}{p_{1}}^{n_{1}}{p_{2}}^{n_{2}}{p_{3}}^{n_{3}}\left( 1-p_{1}-p_{2}-p_{3} \right)^{N-n_{1}-n_{2}-n_{3}}\#\left( 5 \right) \end{aligned}$$

and

$$\begin{aligned} \Pr\left( B | A' \right)=\frac{N!}{n_{1}!n_{2}!n_{3}!(N-n_{1}-n_{2}-n_{3})!}{q_{1}}^{n_{1}}{q_{2}}^{n_{2}}{q_{3}}^{n_{3}}\left( 1-q_{1}-q_{2}-q_{3} \right)^{N-n_{1}-n_{2}-n_{3}}\#\left( 6 \right) \end{aligned}$$

Taking the data from Figure 3B and Table S3 for patients without a germline variant, we have

- $q_{1}=0.0023$ (0.23% probability of single recurrent somatic hotspots)
- $q_{2}=0.0020$ (0.20% probability of single non-recurrent somatic variants)
- $q_{3}=0.0014$ (0.14% probability of multiple somatic variants)

and, from Figure 3E, for patients with the three most prevalent established germline variants (c.3G>A, D140fs, and A500fs), we have

- $p_{1}=0.58$ (58% probability of single recurrent somatic R525H and non-R525H hotspots)
- $p_{2}=0.10$ (10% probability of single non-recurrent somatic variants)
- $p_{3}=0.007$ (0.7% probability of multiple somatic variants)

To evaluate Eq. (2), it remains only to specify the prior probability of pathogenicity $\Pr\left( A \right)$. After computing the posterior, we can calculate the odds of pathogenicity (OddsPath) by rearranging equation 4 from Tavtigian et al. (2018):^8^

$$\begin{aligned} OddsPath=\frac{\Pr\left( A | B \right)(1-\Pr\left( A \right))}{\Pr\left( A \right)(1-\Pr\left( A | B \right))}\#\left( 7 \right) \end{aligned}$$

For example, if we have a sample of nine patients with a particular germline variant under evaluation, three of whom have concurrent somatic hits (say, once with a single R525H, once with a single T227M, and once with a single A346P), then we have $N=9$, $n_{1}=2$, $n_{2}=1$, and $n_{3}=0$. From Eqs. (5) and (6), we calculate that $\Pr\left( B | A \right)=0.008$ and $\Pr\left( B | A^{'} \right)=1.95\times{10}^{-6}$. Specifying a prior probability of pathogenicity $\Pr\left( A \right)=0.1$ (following Tavtigian et al., 2018),^8^ we then have from Eq. (2) that $\Pr\left( A | B \right)=0.998$, i.e., the posterior probability of pathogenicity is 99.8%, and from Eq. (7) the OddsPath are 4087, equivalent to a “very strong” level of evidence for pathogenicity.

**SUPPLEMENTAL TABLES**

**Table S1.** Summary of the *DDX41* aggregated synthetic cohort, including **(A)** 36 study cohorts (from 37 publications) of 53795 consecutive patients, and **(B)** 56 other studies that do not meet the case cohort criteria used in somatic analysis.

**Table S2.** Quasi-case-control study of the association between various germline *DDX41* variants and myelodysplastic syndrome/acute myeloid leukemia (MDS/AML) compared to population controls, according to overall cases and within non-Finnish European (NFE) and East Asian (EAS) genetic ancestry groups. Variants with at least 10 total cases, 3 occurrences within the EAS ancestry group, and/or 5 NFE-only instances are included.

**Table S3.** Contingency table showing the associations between the presence of germline variants (including pathogenic [P], likely pathogenic [LP], and variants of uncertain significance [VUS]) and different types of somatic *DDX41* variants. Odds ratios and 95% confidence intervals are calculated. Posterior probability, based on Meierhofer et al. (2023), is provided for the different observed somatic events.

**Table S4.** Odds of Pathogenicity (OddsPath) based on somatic hits for 239 germline *DDX41* variants. N_total indicates the number of times the germline variant under evaluation is observed. The case numbers of somatic *DDX41* events (single recurrent missense, single non-recurrent, and multiple) are provided according to cohort sources. The evidence strengths based on the modified Maierhofer method and OddsPath are compared; red and blue colors highlight variants upgraded (n=120) and downgraded (n=8) by OddsPath, respectively.

**Table S5.** Confusion matrices illustrating the performance of variant classification across different REVEL scores and AlphaMissense classifications. Each matrix compares the putative variant status (pathogenic [n=61] versus non-pathogenic [n=503], based on the presence of a single recurrent somatic variant) with the predicted classification from each method.

**SUPPLEMENTAl FIGURES**

**Figure S1. Recursive partitioning decision tree of *DDX41* germline variant classification.** Variants are grouped by type: **(A)** start-loss, frameshift, and nonsense variants (n=110); **(B)** canonical splice site and intronic variants (n=54); and **(C)** missense and in-frame variants (n=269). Five missense variants lacking HGVSc information with multiple possible backtranslations were excluded from the analysis. The decision tree is generated based on all ACMG/AMP criteria using the rpart package. The hierarchical splits represent sequential criteria, with higher splits indicating more influential factors in the initial decision-making process. Variants are classified into Pathogenic (P, purple), Likely Pathogenic (LP, blue), or Variant of Uncertain Significance (VUS, gray) based on the algorithm's predictive classification. Each terminal node contains three numbers representing the actual counts of variants with P, LP, and VUS classifications, as described in the Supplemental Methods. Recursive partitioning is a criterion-based analysis for variant type rather than individual variants; therefore, a discrepancy between the exact counts and the assigned pathogenicity class within a terminal node does not indicate a final misclassification but rather reflects the current decision-making state up to that partition.

**Figure S2. Characteristics of germline *DDX41* variants** in 1368 cases of myelodysplastic syndrome and acute myeloid leukemia (MDS/AML) harboring a total of 1378 variants (10 cases have two germline variants). **(A)** Pathogenic and likely pathogenic (P/LP) variants (total n=1089) showing structural and truncating (top), and non-truncating (bottom) variants. **(B)** Less common P/LP truncating variants, a subset of (A), are shown: frameshift/nonsense (top) and splice/intronic (bottom) variants. **(C)** Variants of uncertain significance (VUS) (total n=289) showing missense / in-frame (top) and other (bottom) variants.

**Figure S3.** **Spectrum of *DDX41* variants** **outside the context of myelodysplastic syndrome and acute myeloid leukemia (MDS/AML).** **(A)** Non-MDS/AML myeloid neoplasms and cytopenias, and **(B)** lymphoid neoplasms. The presence of a concurrent somatic *DDX41* variant is indicated with red (yes) or blue (no).

**Figure S4. Summary of single *DDX41* somatic variants** observed alongside a germline *DDX41* variant in 800 myelodysplastic syndrome/acute myeloid leukemia (MDS/AML) cases. One case was excluded due to missing variant information. **(A)** The top seven recurrent somatic variants (top) constituted 674 (84%) of the cases. Rare somatic variants (bottom) included 75 different variants found in 126 (16%) cases; all were missense or in-frame variants except for three: E2* (variant allele fraction [VAF] unknown), S4* (VAF unknown), and c.645-1G>A (VAF 1%), which were all identified alongside the germline R369G variant (in separate cases). **(B)** Violin and dot plot of VAFs among (primarily assumed) somatic *DDX41* variants, showing significant differences between groups (p=2.2e-6 by Kruskal-Wallis test). Pairwise Wilcoxon tests were adjusted using the Benjamini and Hochberg method. P-value annotations: <0.05 (*), <0.01 (**), <0.001 (***), <0.0001 (****).

**Figure S5. Association between less common germline *DDX41* variants and somatic variant types.** The top panel shows germline variants with 5 to 14 occurrences, while the bottom panel displays those with 3 to 4 occurrences.

**Figure S6. Characteristics of somatic-only *DDX41* variants.** The **middle panel** presents a summary of 147 single variants and 97 multiple somatic variants identified in 48 cases (with one case having three variants). The **top** **panel** summarizes the different types of single somatic variants, while the **bottom** **panel** illustrates the relationship between pairs of variants in the 48 cases with double (assumed) somatic-only *DDX41* variants.

**Figure S7. Variant allele fractions (VAF %) of somatic-only *DDX41* variants. (A)** Comparison of VAFs between single somatic *DDX41* variants in cases with and without a germline variant. **(B)** VAFs of single somatic-only *DDX41* variants based on variant types. **(C)** VAFs of double (assumed) somatic-only *DDX41* variants. Variants #1 (circle) and #2 (triangle) represent the higher and lower VAF variants, respectively, and the colors indicate the variant types.

**Figure S8. Comparison of REVEL and AlphaMissense in silico tools.** The ability to classify putative pathogenic (n=61, red) and non-pathogenic (n=503, blue) variants, based on the presence of any concurrent single recurrent somatic variant, was evaluated and compared. **(A)** Density plot and **(B)** histogram of REVEL scores in the variant classification. Dashed vertical lines indicate various REVEL score thresholds. **(C)** Density plot and **(D)** histogram of AlphaMissense in the variant classification.
