## Supplementary material for "Refinement of the Classification of *DDX41* Variants Through Analysis of Aggregated Clinical Datasets": Figure S1

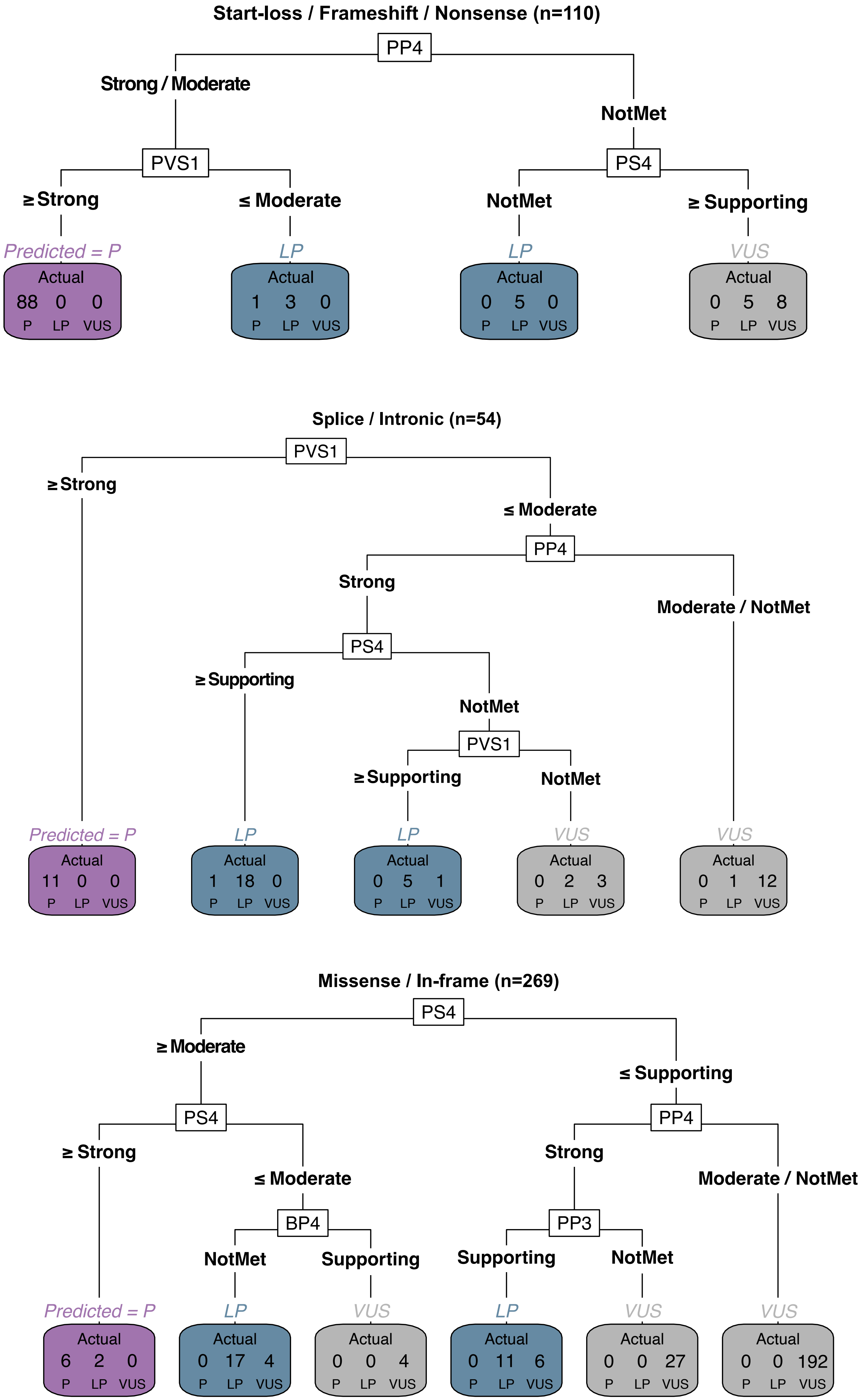

Start-loss / Frameshift / Nonsense (n=110)
PP4
Strong / Moderate
NotMet
PVS1
PS4
≥ Strong
≤ Moderate
NotMet
≥ Supporting
Predicted = P
Actual 88 0 0 P LP VUS
LP
Actual 1 3 0 P LP VUS
LP
Actual 0 5 0 P LP VUS
VUS
Actual 0 5 8 P LP VUS
Splice / Intronic (n=54)
PVS1
≥ Strong
≤ Moderate
PP4
Strong
Moderate / NotMet
PS4
≥ Supporting
NotMet
PVS1
≥ Supporting
NotMet
Predicted = P
Actual 11 0 0 P LP VUS
LP
Actual 1 18 0 P LP VUS
LP
Actual 0 5 1 P LP VUS
VUS
Actual 0 2 3 P LP VUS
VUS
Actual 0 1 12 P LP VUS
Missense / In-frame (n=269)
PS4
≥ Moderate
≤ Supporting
PS4
PP4
≥ Strong
≤ Moderate
Strong
Moderate / NotMet
BP4
PP3
NotMet
Supporting
Supporting
NotMet
Predicted = P
Actual 6 2 0 P LP VUS
LP
Actual 0 17 4 P LP VUS
VUS
Actual 0 0 4 P LP VUS
LP
Actual 0 11 6 P LP VUS
VUS
Actual 0 0 27 P LP VUS
VUS
Actual 0 0 192 P LP VUS

**Figure S1. Recursive partitioning decision tree of *DDX41* germline variant classification.** Variants are grouped by type: **(A)** start-loss, frameshift, and nonsense variants (n=110); **(B)** canonical splice site and intronic variants (n=54); and **(C)** missense and in-frame variants (n=269). Five missense variants lacking HGVSc information with multiple possible backtranslations were excluded from the analysis. The decision tree is generated based on all ACMG/AMP criteria using the rpart package. The hierarchical splits represent sequential criteria, with higher splits indicating more influential factors in the initial decision-making process. Variants are classified into Pathogenic (P, purple), Likely Pathogenic (LP, blue), or Variant of Uncertain Significance (VUS, gray) based on the algorithm's predictive classification. Each terminal node contains three numbers representing the actual counts of variants with P, LP, and VUS classifications, as described in the Supplemental Methods. Recursive partitioning is a criterion-based analysis for variant type rather than individual variants; therefore, a discrepancy between the exact counts and the assigned pathogenicity class within a terminal node does not indicate a final misclassification but rather reflects the current decision-making state up to that partition.
