## Supplementary material for "Refinement of the Classification of *DDX41* Variants Through Analysis of Aggregated Clinical Datasets": Figure S2

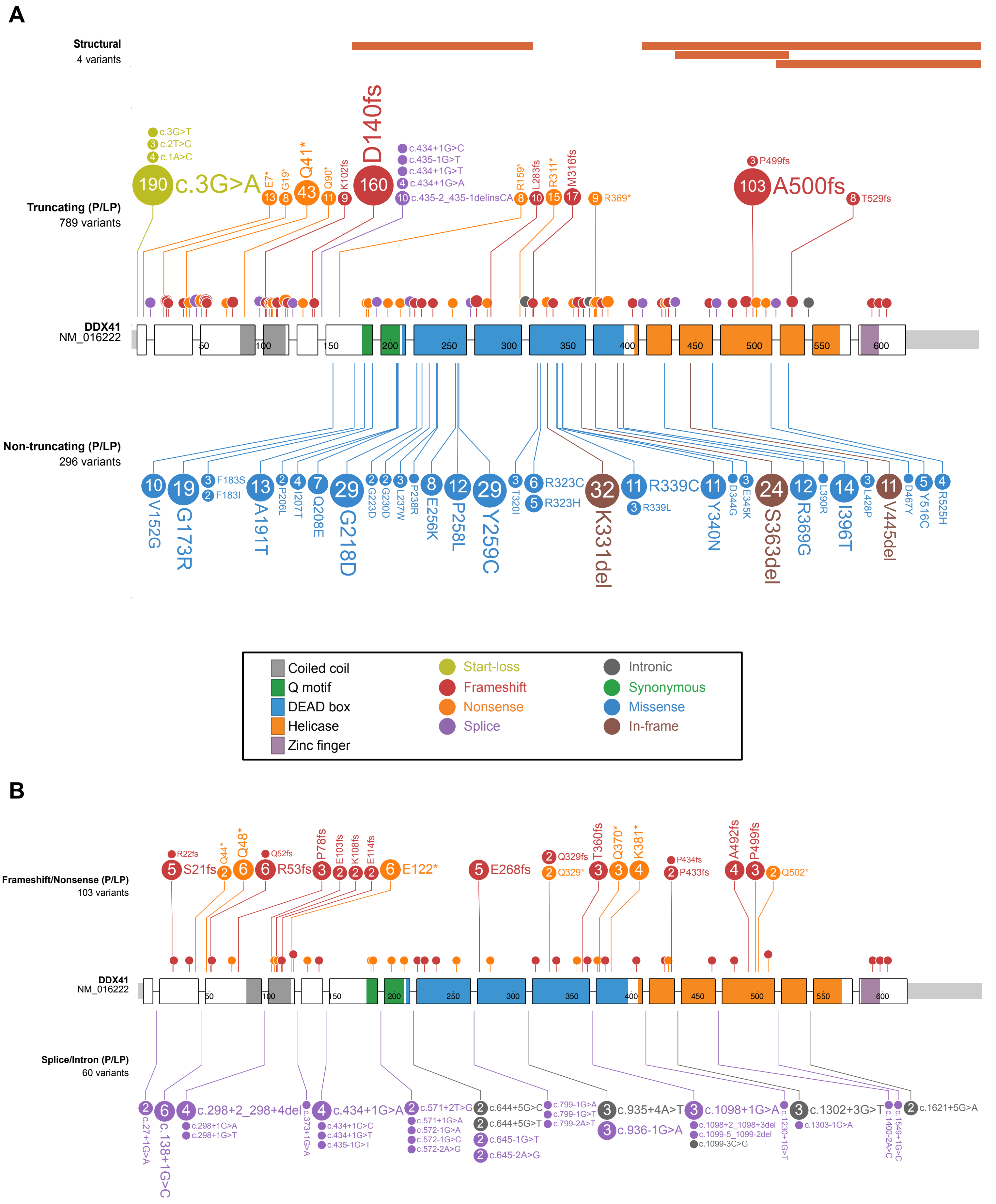
A
Structural
4 variants
Truncating (P/LP)
789 variants
190 c.3G>A
c.3G>T
3 c.2T>C
4 c.1A>C
13 E7*
8 G19*
43 Q41*
11 Q90*
9 K102fs
160 D140fs
c.434+1G>C
c.435-1G>T
c.434+1G>T
4 c.434+1G>A
10 c.435-2_435-1delinsCA
8 R159*
10 L283fs
15 R311*
17 M316fs
9 R369*
3 P499fs
103 A500fs
8 T529fs
DDX41
NM_016222
50
100
150
200
250
300
350
400
450
500
550
600
Non-truncating (P/LP)
296 variants
10 V152G
19 G173R
3 F183S
2 F183I
13 A191T
2 P206L
4 I207T
7 Q208E
29 G218D
2 G223D
2 G230D
3 L237W
P238R
8 E256K
12 P258L
29 Y259C
3 T320I
6 R323C
5 R323H
32 K331del
11 R339C
3 R339L
11 Y340N
D344G
3 E345K
24 S363del
12 R369G
L390R
14 I396T
3 L428P
11 V445del
D467Y
5 Y516C
4 R525H
Coiled coil
Q motif
DEAD box
Helicase
Zinc finger
Start-loss
Frameshift
Nonsense
Splice
Intronic
Synonymous
Missense
In-frame
B
Frameshift/Nonsense (P/LP)
103 variants
R22fs
5 S21fs
2 Q44*
6 Q48*
Q52fs
6 R53fs
3 P78fs
2 E103fs
2 K108fs
2 E114fs
6 E122*
5 E268fs
2 Q329fs
2 Q329*
3 T360fs
3 Q370*
4 K381*
P434fs
2 P433fs
4 A492fs
3 P499fs
2 Q502*
DDX41
NM_016222
Splice/Intron (P/LP)
60 variants
2 c.27+1G>A
6 c.138+1G>C
4 c.298+2_298+4del
c.298+1G>A
c.298+1G>T
c.373+1G>A
4 c.434+1G>A
c.434+1G>C
c.434+1G>T
c.435-1G>T
2 c.571+2T>G
c.571+1G>A
c.572-1G>A
c.572-1G>C
c.572-2A>G
2 c.644+5G>C
2 c.644+5G>T
2 c.645-1G>T
2 c.645-2A>G
c.799-1G>A
c.799-1G>T
c.799-2A>T
3 c.935+4A>T
3 c.936-1G>A
3 c.1098+1G>A
c.1098+2_1098+3del
c.1099-5_1099-2del
c.1099-3C>G
c.1230+1G>T
3 c.1302+3G>T
c.1303-1G>A
c.1400-2A>C
c.1549+1G>C
2 c.1621+5G>A

### Figure S2

C

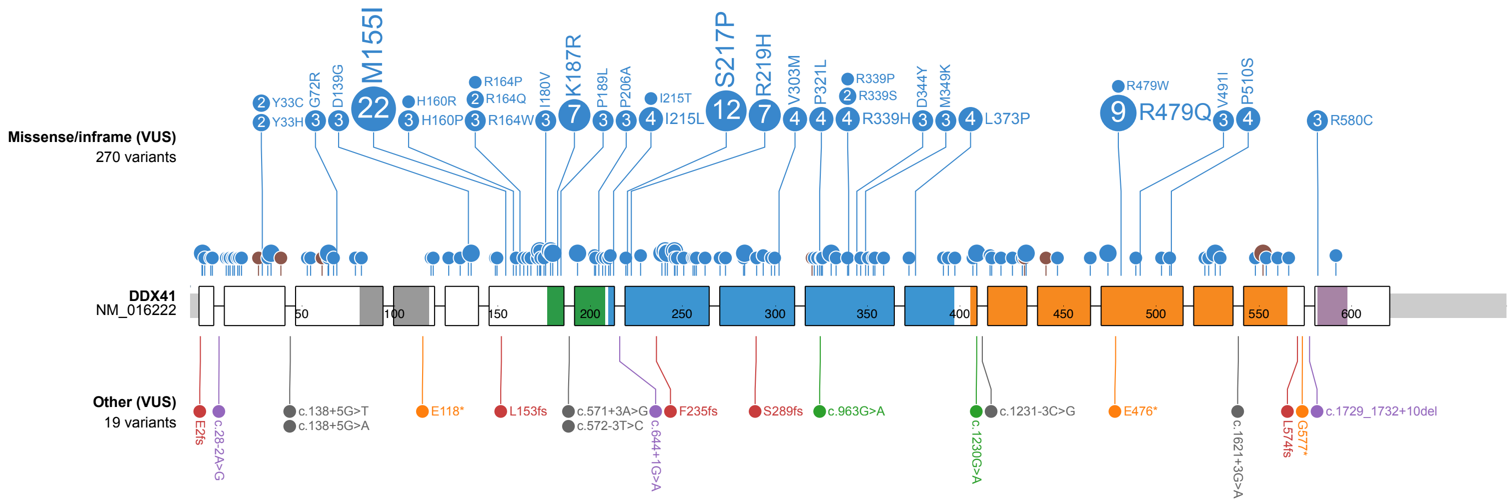


**Figure S2. Characteristics of germline *DDX41* variants** in 1368 cases of myelodysplastic syndrome and acute myeloid leukemia (MDS/AML) harboring a total of 1378 variants (10 cases have two germline variants). **(A)** Pathogenic and likely pathogenic (P/LP) variants (total n=1089) showing structural and truncating (top), and non-truncating (bottom) variants. **(B)** Less common P/LP truncating variants, a subset of (A), are shown: frameshift/nonsense (top) and splice/intronic (bottom) variants. **(C)** Variants of uncertain significance (VUS) (total n=289) showing missense / in-frame (top) and other (bottom) variants.

Missense/inframe (top) and other (bottom) - bottom looks like it includes silent variants and splice region variants that may or may not be truncating.
