## Supplementary material for "Refinement of the Classification of *DDX41* Variants Through Analysis of Aggregated Clinical Datasets": Figure S3

**A**

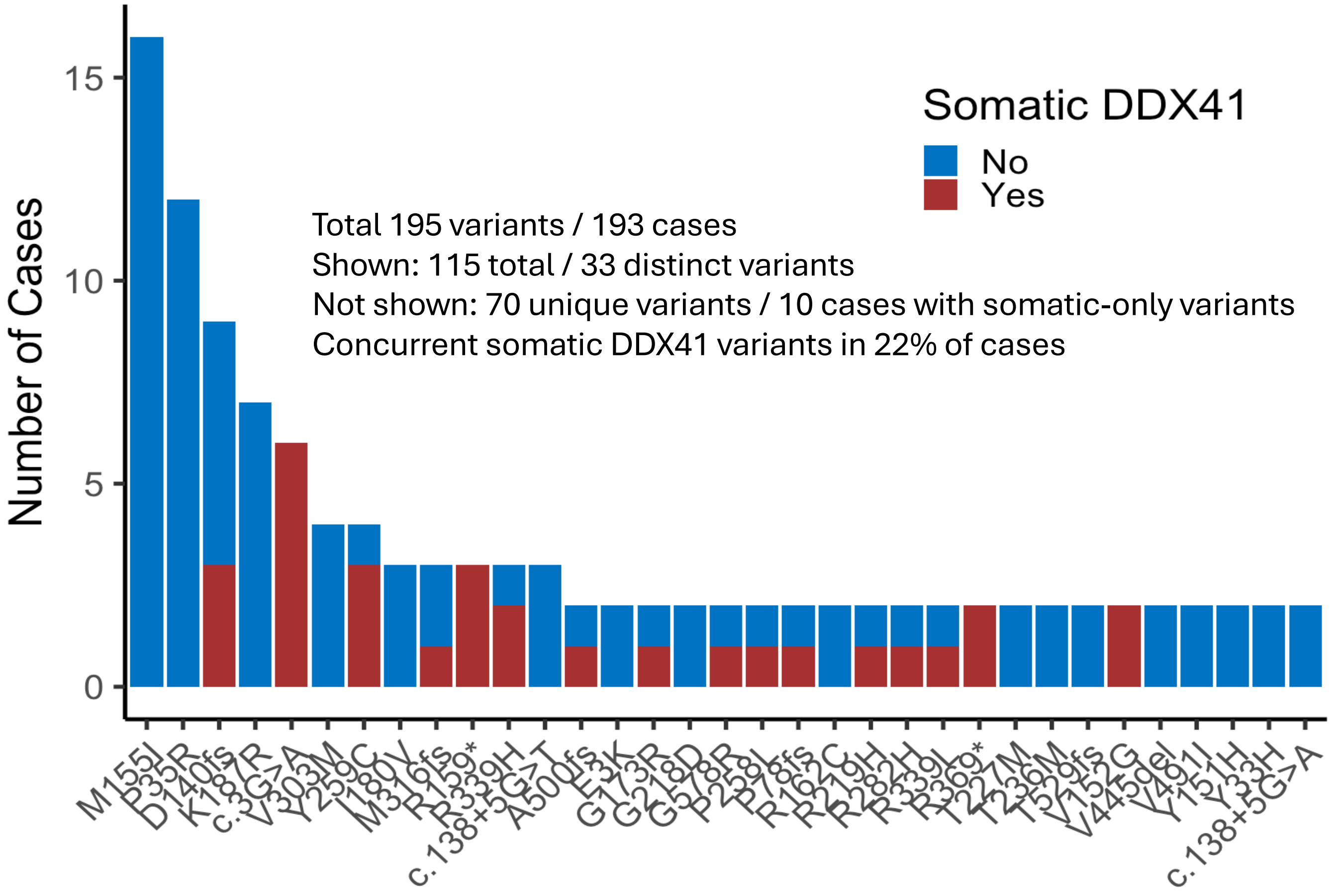


**B**

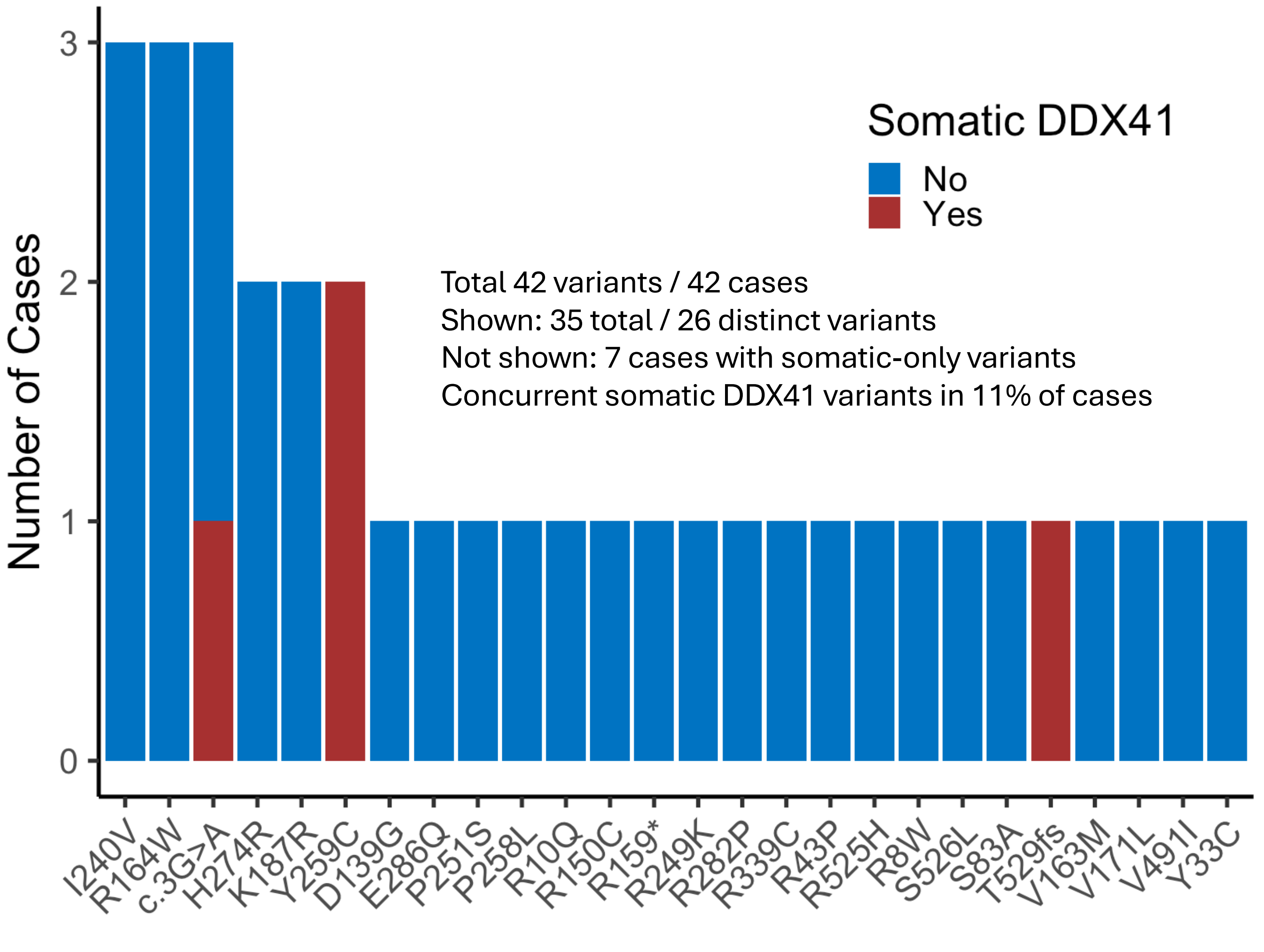


**Figure S3. Spectrum of *DDX41* variants outside the context of myelodysplastic syndrome and acute myeloid leukemia (MDS/AML). (A)** Non-MDS/AML myeloid neoplasms and cytopenias, and **(B)** lymphoid neoplasms. The presence of a concurrent somatic *DDX41* variant is indicated with red (yes) or blue (no).
