## Supplementary material for "Refinement of the Classification of *DDX41* Variants Through Analysis of Aggregated Clinical Datasets": Figure S4

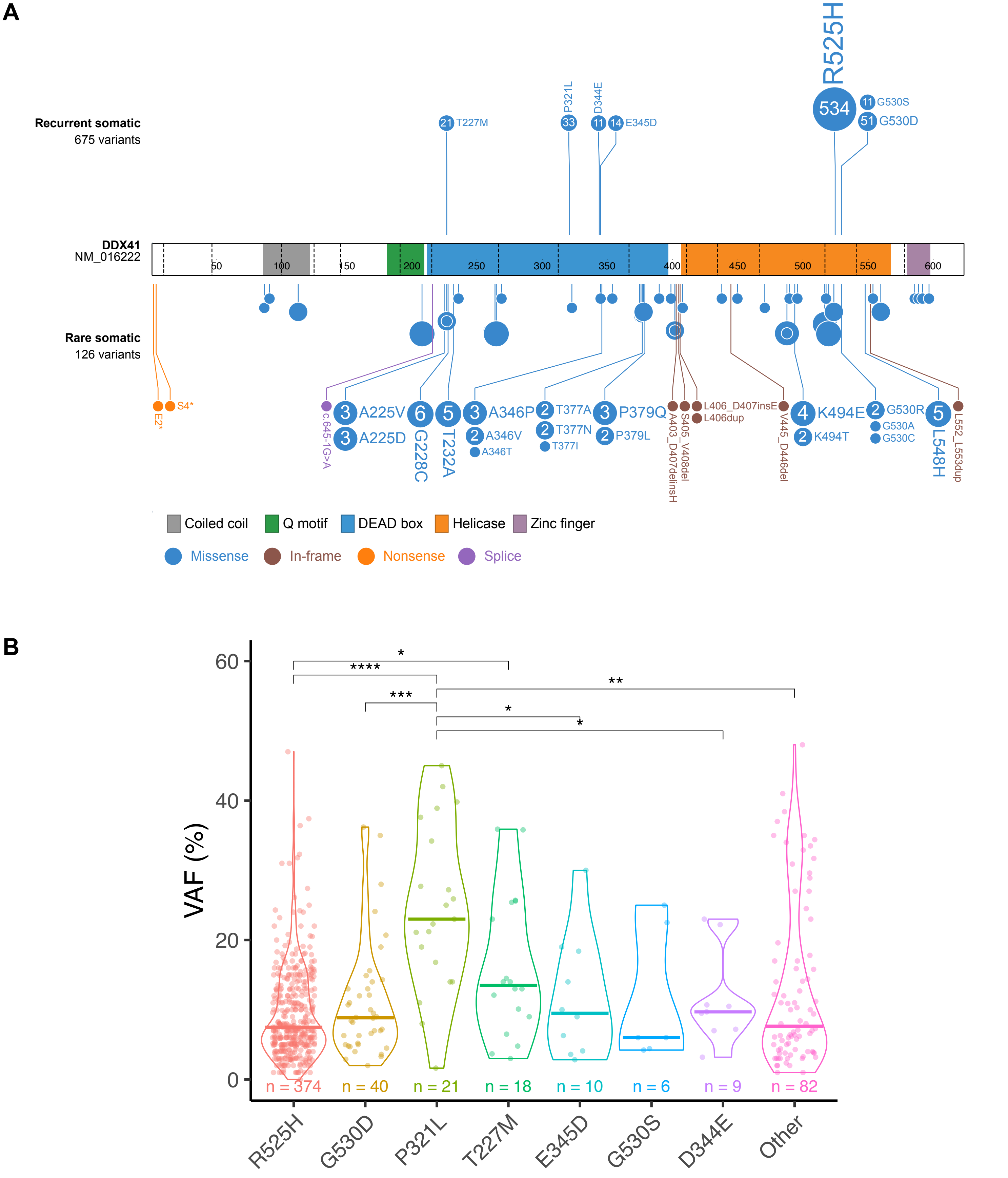


**Figure S4. Summary of single *DDX41* somatic variants** observed alongside a germline *DDX41* variant in 800 myelodysplastic syndrome/acute myeloid leukemia (MDS/AML) cases. One case was excluded due to missing variant information. **(A)** The top seven recurrent somatic variants (top) constituted 674 (84%) of the cases. Rare somatic variants (bottom) included 75 different variants found in 126 (16%) cases; all were missense or in-frame variants except for three: E2* (variant allele fraction [VAF] unknown), S4* (VAF unknown), and c.645-1G>A (VAF 1%), which were all identified alongside the germline R369G variant (in separate cases). **(B)** Violin and dot plot of VAFs among (primarily assumed) somatic *DDX41* variants, showing significant differences between groups (p=2.2e-6 by Kruskal-Wallis test). Pairwise Wilcoxon tests were adjusted using the Benjamini and Hochberg method. P-value annotations: <0.05 (*), <0.01 (**), <0.001 (***), <0.0001 (****).
