## Supplementary material for "Refinement of the Classification of *DDX41* Variants Through Analysis of Aggregated Clinical Datasets": Figure S5

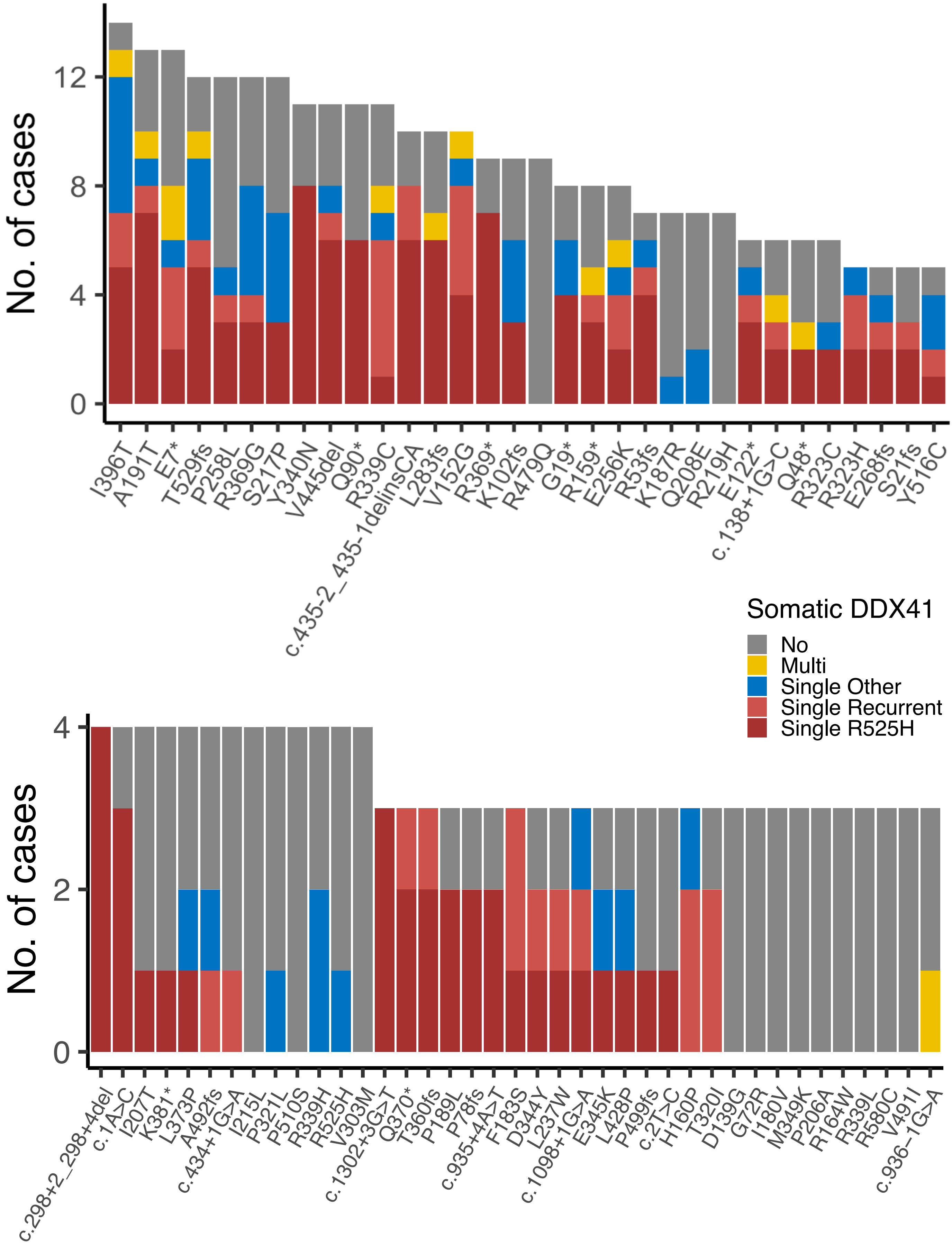


**Figure S5. Association between less common germline *DDX41* variants and somatic variant types.** The top panel shows germline variants with 5 to 14 occurrences, while the bottom panel displays those with 3 to 4 occurrences.
