## Supplementary material for "Refinement of the Classification of *DDX41* Variants Through Analysis of Aggregated Clinical Datasets": Figure S6

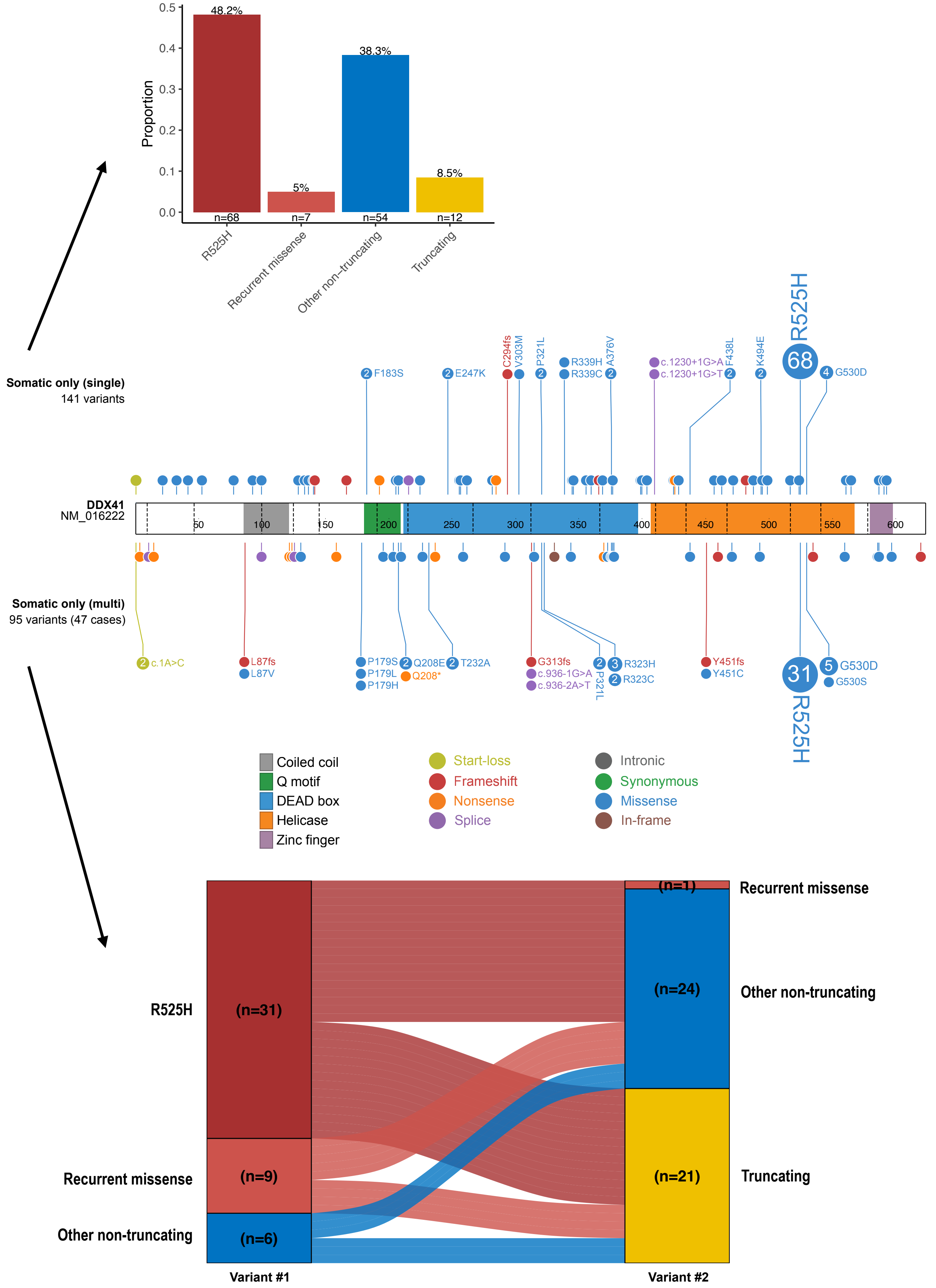


**Figure S6. Characteristics of somatic-only *DDX41* variants.** The **middle panel** presents a summary of 147 single variants and 97 multiple somatic variants identified in 48 cases (with one case having three variants). The **top panel** summarizes the different types of single somatic variants, while the **bottom panel** illustrates the relationship between pairs of variants in the 48 cases with double (assumed) somatic-only *DDX41* variants.
