## Supplementary material for "Refinement of the Classification of *DDX41* Variants Through Analysis of Aggregated Clinical Datasets": Figure S7

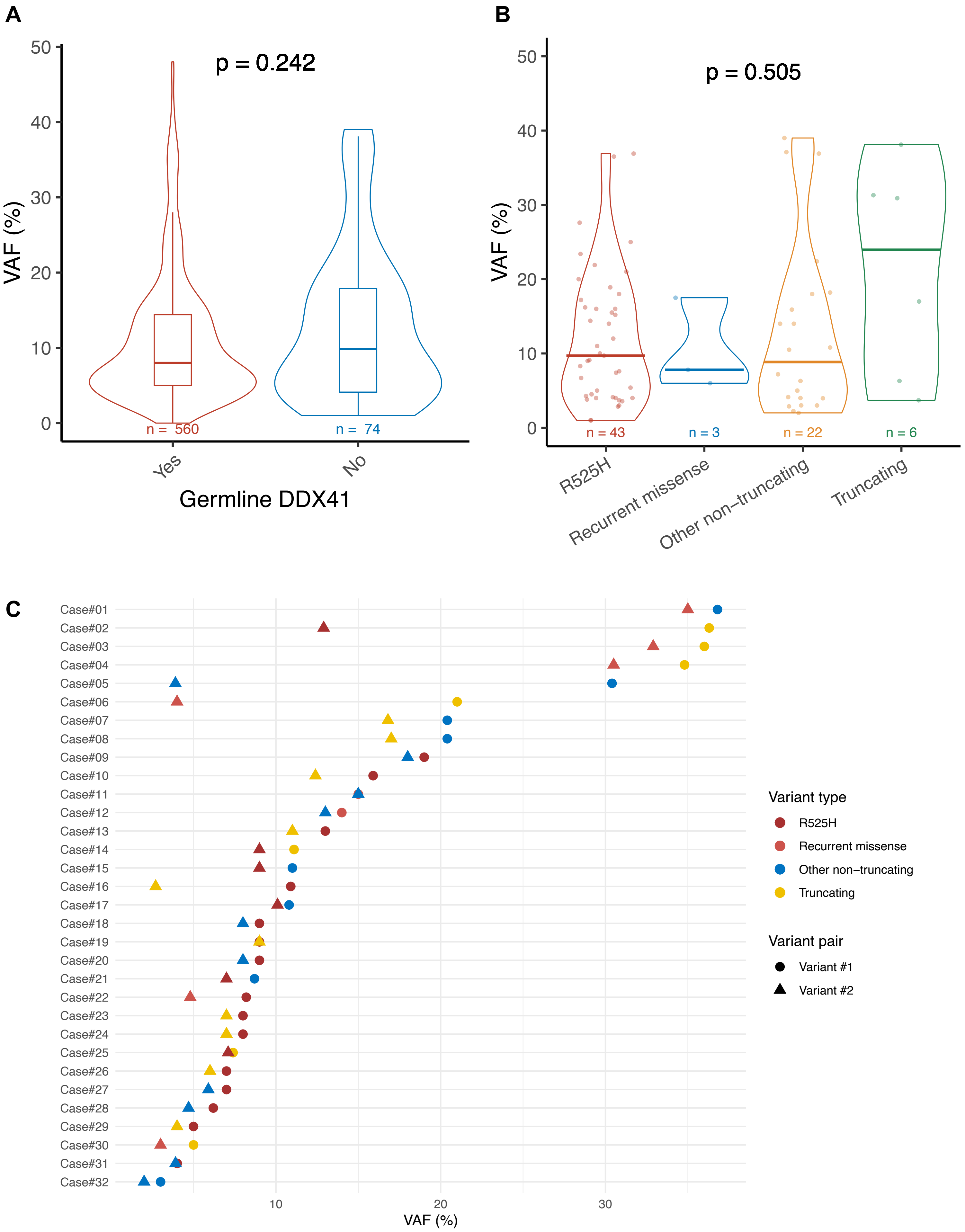


**Figure S7. Variant allele fractions (VAF %) of somatic-only *DDX41* variants. (A)** Comparison of VAFs between single somatic *DDX41* variants in cases with and without a germline variant. **(B)** VAFs of single somatic-only *DDX41* variants based on variant types. **(C)** VAFs of double (assumed) somatic-only *DDX41* variants. Variants #1 (circle) and #2 (triangle) represent the higher and lower VAF variants, respectively, and the colors indicate the variant types.
