## Supplementary material for "Refinement of the Classification of *DDX41* Variants Through Analysis of Aggregated Clinical Datasets": Figure S8

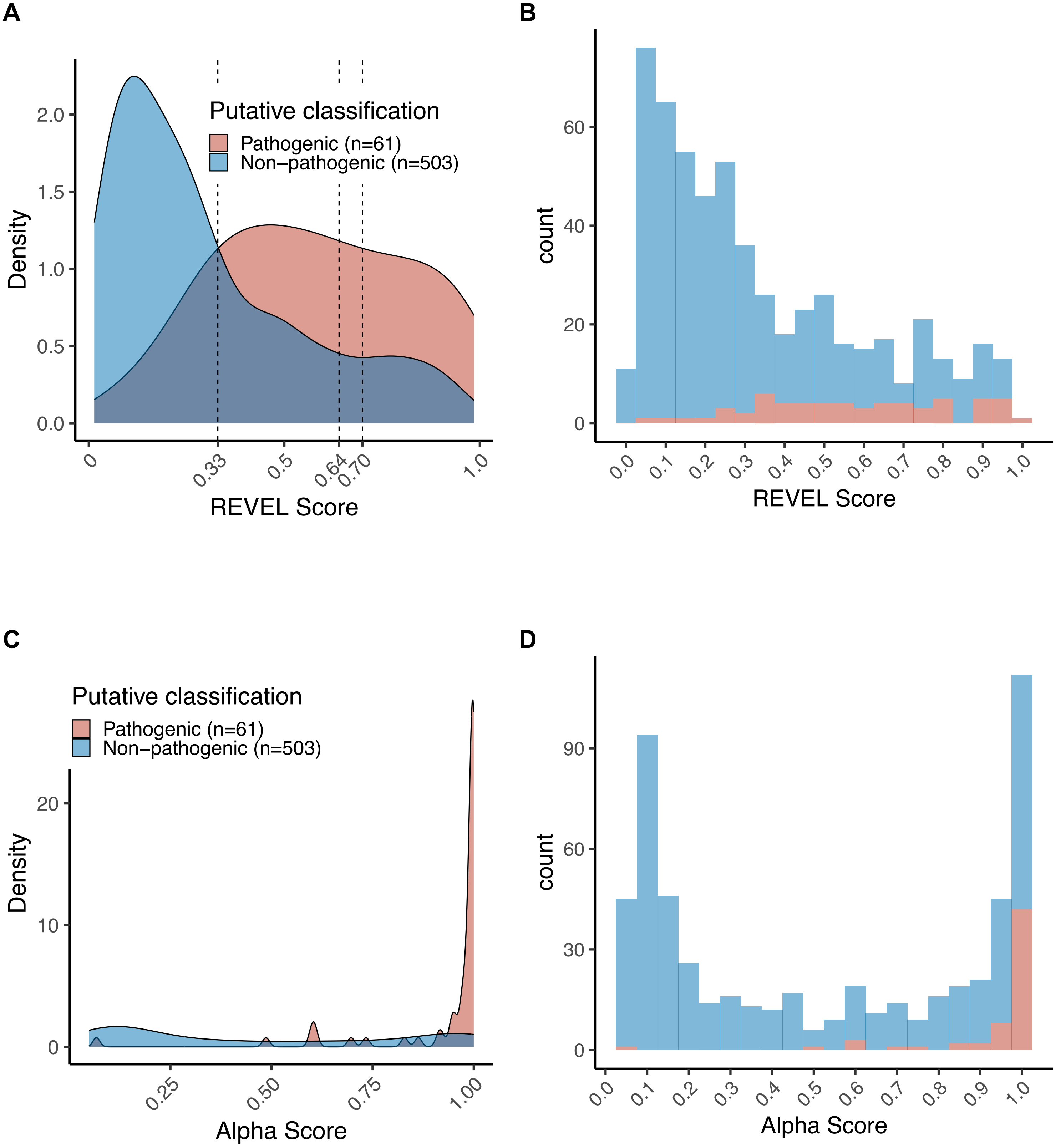


**Figure S8. Comparison of REVEL and AlphaMissense in silico tools.** The ability to classify putative pathogenic (n=61, red) and non-pathogenic (n=503, blue) variants, based on the presence of any concurrent single recurrent somatic variant, was evaluated and compared. **(A)** Density plot and **(B)** histogram of REVEL scores in the variant classification. Dashed vertical lines indicate various REVEL score thresholds. **(C)** Density plot and **(D)** histogram of AlphaMissense in the variant classification.
